# Disproportionate incidence of COVID-19 in African Americans correlates with dynamic segregation

**DOI:** 10.1101/2020.07.08.20148742

**Authors:** Aleix Bassolas, Sandro Sousa, Vincenzo Nicosia

## Abstract

Socio-economic disparities quite often have a central role in the unfolding of large-scale catastrophic events. One of the most concerning aspects of the ongoing COVID-19 pandemics [1] is that it disproportionately affects people from Black and African American backgrounds [2–6], creating an unexpected infection gap. Interestingly, the abnormal impact on these ethnic groups seem to be almost uncorrelated with other risk factors, including co-morbidity, poverty, level of education, access to healthcare, residential segregation, and response to cures [7–11]. A proposed explanation for the observed incidence gap is that people from African American backgrounds are more often employed in low-income service jobs, and are thus more exposed to infection through face-to-face contacts [12], but the lack of direct data has not allowed to draw strong conclusions in this sense so far. Here we introduce the concept of dynamic segregation, that is the extent to which a given group of people is internally clustered or exposed to other groups, as a result of mobility and commuting habits. By analysing census and mobility data on more than 120 major US cities, we found that the dynamic segregation of African American communities is significantly associated with the weekly excess COVID-19 incidence and mortality in those communities. The results confirm that knowing where people commute to, rather than where they live, is much more relevant for disease modelling.

---

The spread of a non-air-borne virus like COVID-19 is mostly mediated by direct face-to-face contacts with other infected people. This is why the first measures attempting at containing the spread of the virus included the introduction of travel restrictions, social distancing, curfews, and stay-at-home orders [13–16]. However, the distribution of the number of contacts per person is known to be fat-tailed [17], so that most of the infections are actually caused by a relatively small set of individuals, called *super-spreaders* [18, 19], who normally have a disproportionately high number of face-to-face contacts. Intuitively enough, super-spreaders are most commonly found among service workers –cashiers, postmen, clerks, cooks, bus drivers, waiters, etc.– since their job involves being in direct contact with a large number of people on a regular basis. This fact makes super-spreaders more prone to catch diseases that propagate preferentially through direct contacts, like COVID-19 does, and –involuntarily– more efficient at spreading them.

The fact that mainly African Americans seem to be affected by such a markedly unusual COVID-19 incidence[20–22], rather than, say, people with lowincome, little access to healthcare, or with other increased risk factors [7–11], points to ethnic segregation, i.e., the tendency of people belonging to the same ethnic group to live closer in space, as a possible culprit [23–27]. Indeed, ethnic segregation is long-standing problem across the US [28], so the idea that the abnormal proportion of COVID-19 infections among African Americans could be due to spatial segregation does not sound unreasonable. However, the results available so far confirm that, although there is a correlation between ethnic segregation and overall incidence of COVID-19 in the population, there seems to be little evidence of an association with infection gap in African Americans [29].

Our hypothesis is that the observed infection gap is most probably due to a prevalence of *super-spreading behaviours* in African American communities, i.e., activities that contribute to increase the typical number and variety of face-to-face contacts of individuals —including for instance their job, habits, social life, commuting and mobility patterns— and that effectively make them more exposed to the infection. In particular, we argue that these super-spreading behaviours are connected to the presence of what we call *dynamic segregation*. By dynamic segregation we mean the extent to which individuals of a certain class or group are either preferentially exposed to other groups, or internally clustered, as a result of their mobility patterns. In this sense, dynamic segregation is somehow complementary to the classical notion of segregation based on residential data, and is instead related with similar measures of segregation based on the concept of activity space [30]. In principle, the fact that a certain residential neighbourhood has an overabundance of people belonging to a single ethnic group might have *per se* little or no role in increasing the probability that those people catch COVID-19. Conversely, the fact that a group of people works preferentially in specific sectors, or in specific areas of a city, almost automatically increases the typical number of face-to-face contacts they have during a day, e.g., by forcing them to commute long distances in packed public transport services.

## RESULTS

### A. Model

We quantify the dynamic segregation of a certain group in a urban area by means of the typical time needed by individuals of that group to get in touch with individuals of other groups when they move around the city. In our model, a city is represented by a graph 𝒢 where nodes are census tracts and each edge indicates a relation between two areas, namely either physical adjacency or the existence of commuting flows between them. Each node is assigned to a class, according to the ethnicity distribution in the corresponding area (see Methods for details). Then, we consider a random walk on the graph 𝒢, and we look at the statistics of Class Mean First Passage Times (CMFPT) and Class Coverage Times (CCT). The former is the number of steps needed to a walker starting on a node of a certain class *α* to end up for the first time on a node of class *β*, while the latter is related to the time needed to a random walk to visit all the classes in the system (see Methods for details). The underlying idea is that a random walk through the graph preserves most of the information about correlations and heterogeneity of node classes [31]. Consequently, if a system is dynamically segregated, the statistics of CMFPT and CCT will be substantially different from those observed on a nullmodel graph having exactly the same set of nodes and edges, but where a node is assigned a class at random from the underlying ethnicity distribution.

In Fig. 1 we provide a visual sketch of the model and we show the distributions of CMFPT and CCT in Chicago and Los Angeles. We chose these two specific cities since Illinois and California are two states respectively characterised by a relatively high and a relatively low incidence gap [32, 33] (a detail of incidence gap across US states is available in Supplementary Figures 18-19). Here each node is associated to one of the seven high-level ethnic groups defined by the US Census Borough [34], with a probability proportional to the abundance of that ethnicity in the corresponding census tract. The variables of interest are 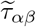 and 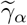. These are, respectively, the ratio of the CMFPT from class *α* to class *β* in the real system and in the null-model, and the ratio of the CCT when the walker starts from class *α* in the real system and in the null model (see Eq. 8 and Eq. 10 in Methods). In short, the farther away 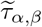 is from 1, the higher the dynamic segregation from class *α* to class *β*. Similarly, the higher the value of 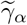 the more isolated ethnicity *α* is from all the other ones.

**FIG. 1.**
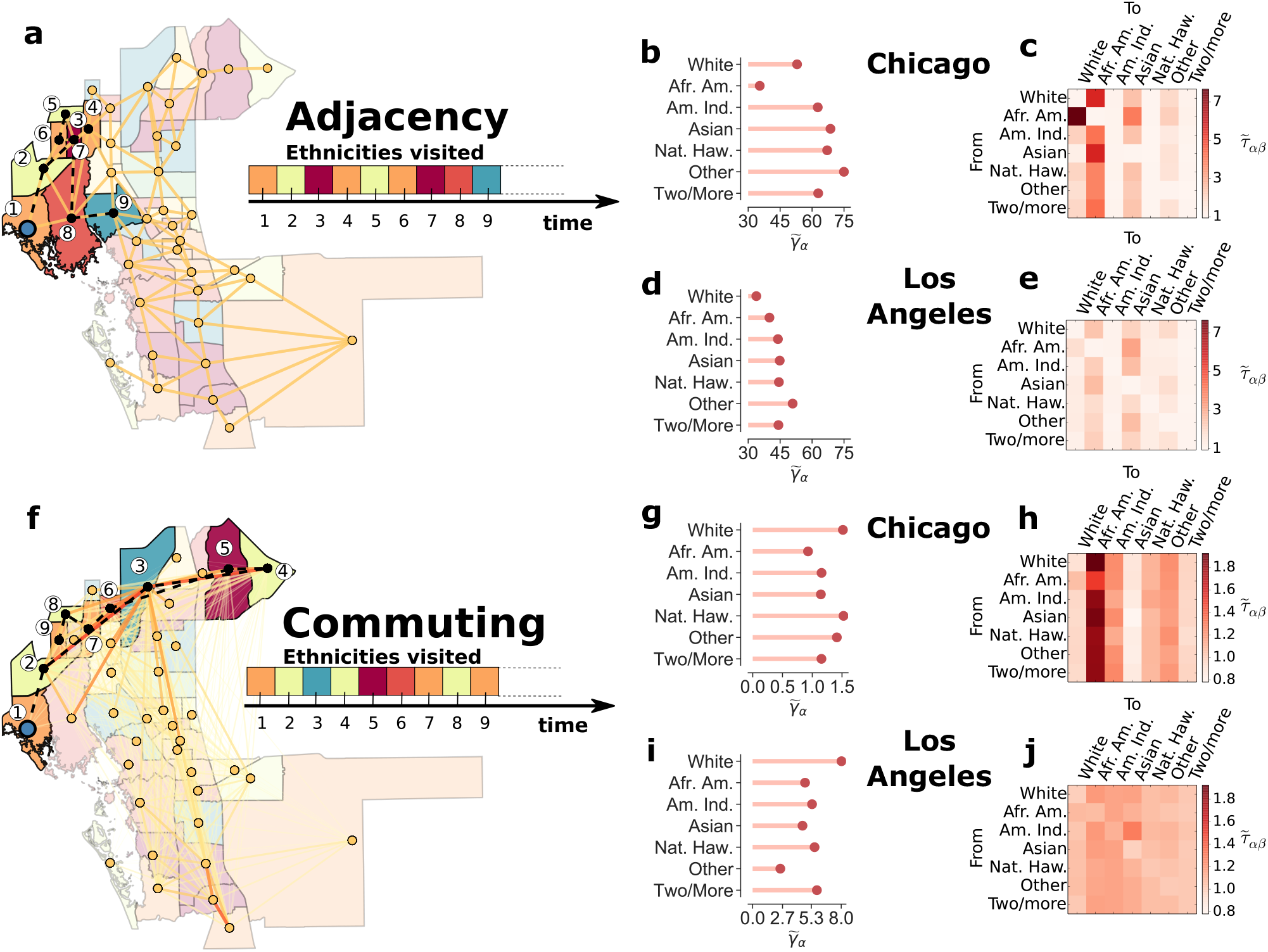
Using typical times of random walks to quantify urban dynamic segregation. The sequence of ethnicities (here indicated by different colours) visited by a random walk over **a** the adjacency network or **f** the commuting network among census tracts of a city retains relevant information about the presence of spatial correlations in ethnicity distribution. Indeed, the normalised values of Class Coverage Time 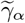 (panels **b**,**d**,**g**,**i**) and Class Mean First Passage Time 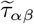 (panels **c**,**e**,**h**,**j** of a random walk exhibit different patterns in different cities, and reveal different kinds of ethnic correlations in the adjacency and in the commuting network of the same city. We show here the values for Chicago or Los Angeles, since Illinois and California have, respectively, one of the highest and one of the lowest COVID-19 incidence gap. Indeed, the mean first passage time from African American to White neighbourhoods in the adjacency graph is much higher in Chicago than in Los Angeles, while the commuting graphs reveals that African Americans are much more exposed to all the other ethnicities in Chicago than in Los Angeles.

The top panels of Fig. 1 correspond to the unweighted network 𝒜 of physical adjacency between census tracts, while the bottom panels are obtained on the weighted network 𝒞 of typical daily commute flows among the same set of census tracts [35] (see Methods for details). Notice that the two graphs have quite different structures: the adjacency graph is planar and each edge connects only nodes that are physically close, while in the commuting graph long edges between physically separated tracts are not only possible, but quite frequent. As a consequence, the adjacency graph provides information about short trips, e.g., for daily shopping and access to local services, while the commuting graph represents long-range trips, e.g., related to commuting to and from work. It is clear that each ethnicity has a peculiar pattern of passage times to the other ethnicities, and this pattern varies across cities. For instance, in Chicago the two largest values of 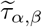 on the adjacency graph are observed between African Americans and White, and between Asian and African Americans. Conversely, in Los Angeles the two largest values of 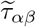 are between African American and Asian and between Other and Asian. As expected, the profile of 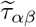 for a given class is quite different if we consider the commuting network instead of the adjacency graph. In Chicago, the largest value of 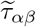 is from White to African American, while in Los Angeles there are a lot of pairs of classes with pretty similar values of 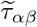, indicating that in this city dynamic segregation for African Americans is less prominent than in Chicago. The value of 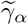 for African Americans is especially low in Chicago, but noticeably different from that of the other ethnicities in Los Angeles. Results for other cities are discussed in Appendix A Supplementary Figures 1-4. As we shall see in a moment, 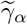 is related to the isolation of a class, so that lower values correspond to increased exposure to all the other classes.

### B. Dynamic segregation and infection gap

Starting from the statistics of CMFPT and CCT at the level of each city, we defined three indices of dynamic segregation, namely dynamic clustering (C), dynamic exposure (E), and dynamic isolation (I), and we associated to each state in the US the weighted average of each of those indices across the largest metropolitan areas of the state (the definitions of these measures are provided in Methods, while a ranking of US states by each segregation index is reported in Appendix B and in Supplementary Figure 5). We considered two temporal data sets of weekly percentage of African Americans infected by and deceased due to COVID-19 for each state in the US [32, 33] (more details available in Methods), and we calculated the incidence gap Δ*A*_inf_ in each state as the difference between the percentage of infected of that state that are African Americans and the percentage of African American population in the same state. Hence, Positive values of Δ*A*_inf_ correspond to a disproportionate incidence of COVID-19 on African American communities.

In Fig. 2 we show the scatter plots of the average dynamic clustering, exposure, and isolation of African Americans at state level, and of the corresponding COVID-19 infection gap in the first two weeks after major lock-down measures were introduced across the US. We chose these two temporal snapshots because the number of confirmed infected individuals in a week actually depends on their contacts up to two weeks before, due to the COVID-19 incubation period [36]. The top panels report the results on the adjacency networks of census tracts, while the bottom panels are for the commuting graphs. Interestingly, there exists a quite strong correlation between dynamic segregation and the disproportionate number of infected in African American communities. In particular, the dynamic clustering of African Americans in a state correlates positively and quite strongly with the infection gap observed in that state in the first two weeks of the data set, both on the adjacency (respectively *R*^2^ = 0.58 and *R*^2^ = 0.44 in the first two weeks) and in the commuting network (respectively *R*^2^ = 0.60 and *R*^2^ = 0.50). This means that if African American citizens normally require more time than citizens from other ethnic groups before ending up in a non-African American neighbourhood, then the incidence gap will be considerably higher.

**FIG. 2.**
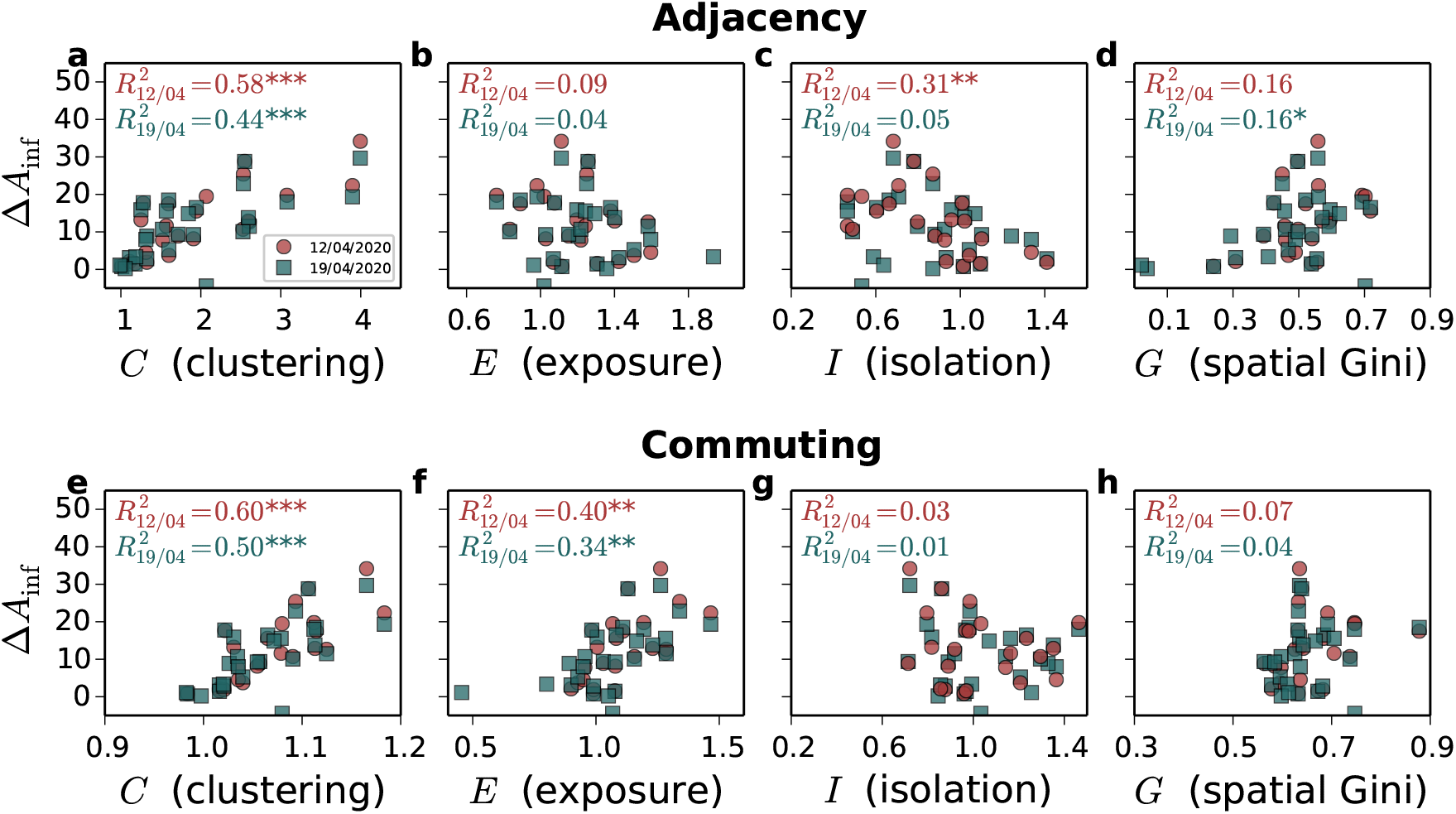
Correlation between incidence gap and dynamic segregation in the early stages of the epidemics. The incidence gap Δ*A*_inf_ across US states in the first two weeks after extensive lock-down measures were enforced exhibits somehow strong correlation with measure of segregation based on CMFPT and CCT on the adjacency (panels **a-d**) and on the commuting graphs (panels **e-h**). In particular, the dynamic clustering *C* (**a**,**e**) is always positively correlated with Δ*A*_inf_, the the dynamic exposure *E* (**b**,**f**) is positively correlated with Δ*A*_inf_ only in the commuting network, and the dynamic isolation *I* (**c**,**g**) is negatively associated with incidence gap only in the adjacency network. Notice that classical measures of residential segregation, like the Spatial Gini coefficient (**d**,**h**), are instead poorly or not correlated at all with incidence gap. Each colour corresponds to a temporal snapshot of the data set, red for 12*/*04*/*2020 and blue for 19*/*04*/*2020. (*: *p <* 0.05,**: *p <* 0.01, ***: *p <* 0.001)

The role of dynamic exposure is even more interesting. Indeed, the dynamic exposure on the adjacency network is not correlated at all with incidence gap, while it is a good predictor of incidence gap in the commuting graph (respectively *R*^2^ = 0.40 and *R*^2^ = 0.36). Conversely, the dynamic isolation of African Americans in the adjacency graph is negatively correlated with incidence gap in the early stages of the epidemics (*R*^2^ = 0.31). Similar results are obtained when we consider the correlation with the death gap Δ*A*_dec_, and the ratios of infection/deaths incidence instead of the difference (see Supplementary Figures 8-10). In particular, dynamic isolation exhibits a somehow stronger correlation with death gap (*R*^2^ = 0.27). It is worth noting that the isolation of other ethnicities has poor or no correlation with incidence gap (see Supplementary Figures 11-14).

The fact that ethnic segregation does not correlate with infection gap as much as dynamic segregation indices can bet better explained by looking at how residential data and dynamic segregation are distributed across a city. In Fig. 3 we show the heat-maps of abundance of African American residents in Chicago and Los Angeles together with the local segregation indices 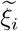 and 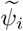, respectively derived from passage times and coverage times, on the adjacency and on the commuting graph of census tracts (see the definitions provided in Methods, additional maps for Detroit and Houston are reported in Appendix C and Supplementary Figure 15). It is true that in Chicago 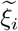 in the adjacency graph is still somehow correlated with the fraction of African American population (see Supplementary Figure 16). But the distribution of 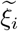 in the commuting graph is totally different. In particular, the regions characterised by residential clusters of African Americans exhibit lower values of 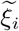, meaning that the commuting patterns make those neighbourhoods overall less isolated. Conversely, new hot-spots are identified in the South-Eastern region of Gary, likely due to the fact that people in this region do not commute much to the city centre anyway. Similarly, the areas of Los Angeles with the largest local isolation are not the neighbourhoods with a higher percentage of African Americans residents, rather the suburbs characterised by high commuting.

**FIG. 3.**
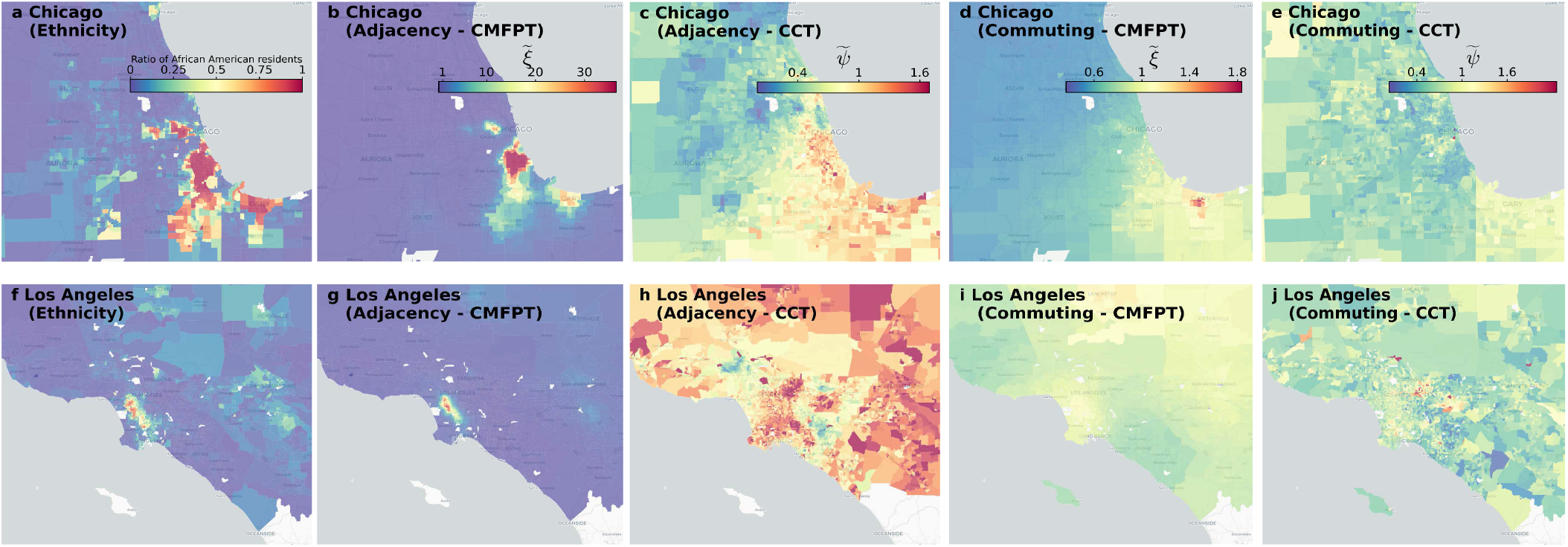
Distribution of local dynamic segregation. The distribution of the fraction of African American population living in each census tract (panels **a**,**f**) is mostly unrelated to the local clustering index 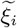 (panels **b**,**d**,**g**,**i**) and to the local isolation index 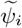 (panels **c**,**e**,**h**,**j**). The figure shows the result for Chicago (top panels) and for Los Angeles (bottom panels). Overall, there is little correlation between the density of African American residents and the dynamic segregation of African Americans in an area. This explains why dynamic segregation indices in a city correlate quite strongly with the COVID-19 infection gap, while no strong association with residential segregation has been found so far.

### C. Combined effects of dynamic segregation and use of public transport

Finally, in Fig. 4**a-d** we show the correlation between the infection gap and the different segregation measures as the pandemic progresses. Unsurprisingly, the correlation with any single measure decreases over time for all the indices, and both on the adjacency and on the commuting graph. Similar results are found for the correlation with death gap and with ratios of incidence and deaths in African Americans (see Appendix D and Supplementary Figures 20-22) as well as with a second dataset we had access to [33] (see Supplementary Figures 23-26). The main reason for the observed decreases is that once large-scale mobility restrictions are put in place —as it happened between the end of March and the beginning of April across all the US states with stayat-home orders and curfews— the overall mobility structure of each city is massively disrupted. As a result, super-spreading behaviours due to usual commuting patterns are massively reduced, and the contagion progresses mainly through face-to-face interactions happening close to the residential place of each individual, and are not captured well by CMFPT and CCT on the commuting graph.

**FIG. 4.**
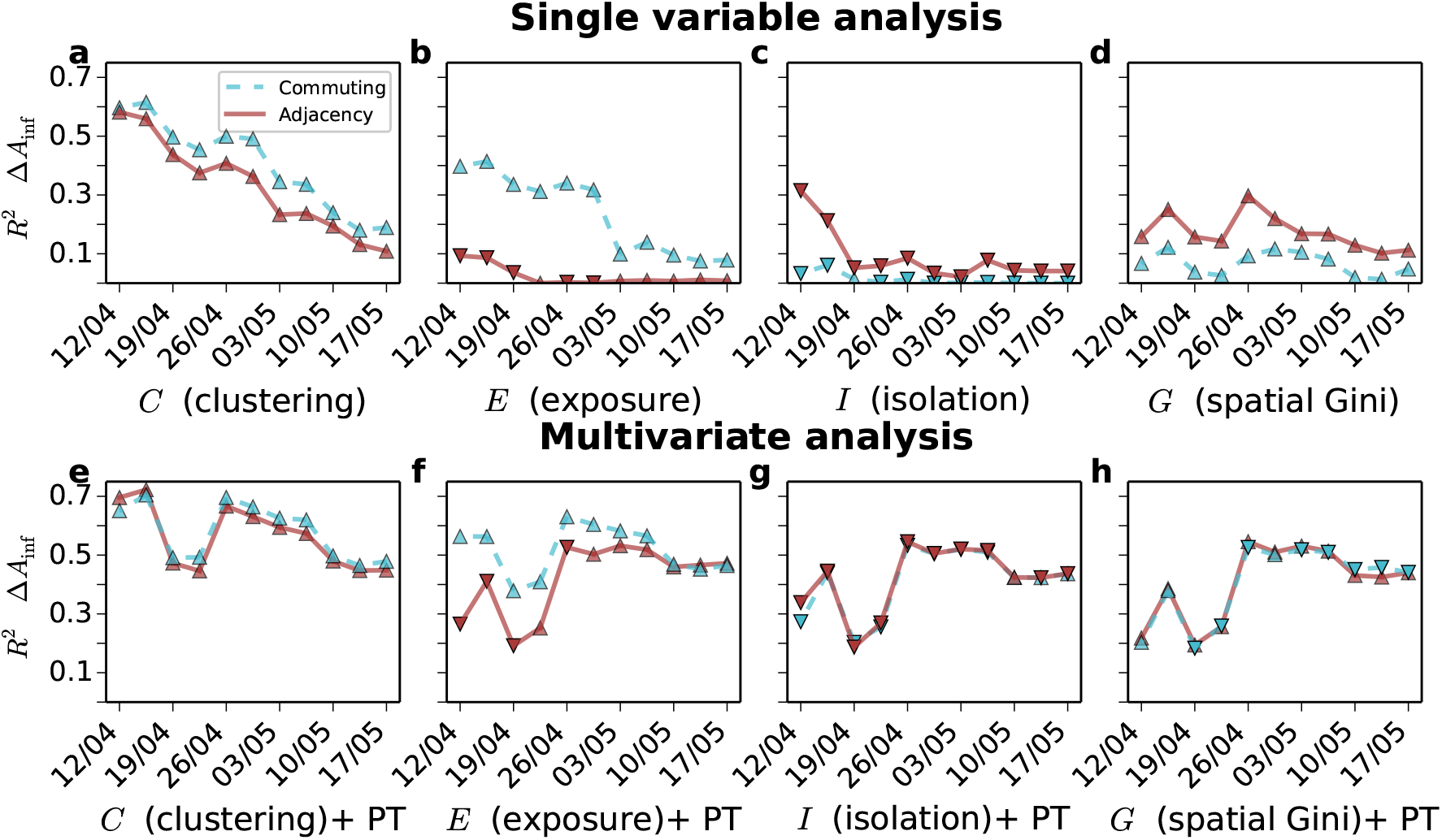
Temporal evolution of incidence gap correlations and multivariate analysis with public transport usage. Evolution of the Pearson correlation (*R*^2^) between African American incidence gap and **a** dynamic clustering, **b** dynamic exposure, **c** dynamic isolation, **d** Spatial Gini coefficient, respectively on the adjacency (solid red lines) and commuting graphs (dashed blue lines). **e-h** Multivariate analysis of the same indices and usage of public transportation by African Americans for **e** dynamic clustering, **f** dynamic exposure, **g** dynamic isolation, **h** and spatial Gini coefficient. The type of marker indicates the sign of the correlation (triangles pointing up for positive correlations, and down for negative correlation). Given the uneven temporal reporting of ethnicity data, each temporal snapshot has a slightly different number of US states (details provided in Supplementary Figure 17). We have also tried alternative formulations for *C* and *E* obtaining significant correlations, as shown in Supplementary Figure 27.

In order to capture the focus on local transport after lock-downs are enforced, in Fig. 4**e-h** we show the results of the multivariate analysis of the same set of segregation indices shown in Fig. 2 and of the fraction of African American population using public transport in each city (see Methods for details). The combination of dynamic segregation and use of public transport correlates quite consistently with the incidence gap. These findings are made more relevant by the fact that the incidence gap in African Americans in the same period is quite poorly correlated with the overall usage of public transport in the population, as well as with a variety of other socioeconomic indices, as shown in Supplementary Figures 30-31. Since cities are complex interconnected systems, it is plausible to hypothesise that segregation and public transport usage are related in subtle and intricate ways, so that it is practically impossible to establish whether the former has caused the latter, or instead the two phenomena have co-evolved over time.

## DISCUSSION

The vulnerability of African American communities and their higher socio-economic disparities has been a standing issue in the US long before the pandemic, the disproportional infection rates simple highlighted and amplified the problem. The presence of a COVID-19 incidence gap in Black and African American population is somehow unexpected, since no specific biological risk factor has been strongly associated with an increased vulnerability to the virus of any specific ethnic group. Hence, the most unbiased assumption to explain such a disproportionate incidence, which in some areas is three to five times higher than the fraction of African American population, is that it should be related to behavioural and social factors, rather than to biological ones. The most frequently whispered theory is that African Americans are more exposed to COVID-19 because they are more frequently employed in service works. This explanation is indeed reasonable, since service workers normally have hundreds of face-to-face interactions during a day. Indeed, some recent studies have estimated that the switching to remote-working was mainly available to people employed in non-essential services, and amounted to 22%-25% of the work force before April [37]. As expected, service workers are one of those categories to which the option to switching to remote-working during the lock-down was not available at all, especially in sectors deemed vital for the functioning of a country during lock-down, including food production and retailers, healthcare, transportation, and logistics. According to the US Labor Force Statistics [38], the occupations with the highest concentration of African Americans are indeed jobs characterised by face-to-face interaction, and most of them fall in the area of *essential* jobs: postal service sorters/processors (42%), nursing (37%), postal service clerks (35%), protective service workers (34%) and barbers (32%). It would not then come as a surprise to discover that one of the major early COVID-19 outbreaks happened in South Dakota, in a meat-processing plant, whose workers were mainly of African American background [39].

The potential relation between ethnicity and mobility was somehow hinted to in a recent study [40] which found that the decrease in the usage of subway transport in New York during the lock-down was uneven across ethnicities, with African Americans experiencing the smallest relative drop. But unfortunately, the publicly available data about COVID-19 incidence do not contain detailed information about socio-economic characteristics of infected individuals, so drawing an association between African Americans, employment in essential service jobs, availability of remote-working options, and increased COVID-19 exposure is very hard.

An interesting finding of the present work is that the combination of dynamic segregation and use of public transport seems to explain the persistence of infection gap throughout the early phases of the pandemic. Indeed, before lock-downs are put in place, African Americans are found to be more exposed to the virus, mainly due to the structure of their daily commuting patterns. After lock-downs are enforced, instead, they are more likely to pass the virus over to other African Americans, as a result of the high levels of clustering and isolation of these communities measured in the adjacency graphs of census tracts, which are a more reliable proxy for face-to-face interactions when long-distance commuting is disrupted. In general, the states where African Americans are more exposed with respect to long-distance trips are also those where they are more clustered with respect to short-range mobility (the rank correlation between the two measures is 0.62, as shown in Supplementary Figure 6-7 and Supplementary Table I).

The importance of considering the interaction of different classes due to mobility through the urbanscape has recently received some attention [30, 41–46]. In this sense, it is quite interesting that the simple diffusion model we used here to quantify the presence of dynamic segregation, and the corresponding indices of clustering, exposure, and isolation, are able to unveil a relatively strong correlation between the structure of mobility in a metropolitan area and the excess incidence of COVID-19 infections and deaths in African Americans. Although the model we consider uses relatively small and coarsegrained information about a city —placement of census tracts, local ethnicity distribution, and commuting trips among them— the strong correlation between dynamic segregation and incidence gap allows to conclude that when it comes to predicting the exposure of a group to a non-airborne virus, knowing the places where the members of that group commute for work is more important and more relevant than knowing where they actually live. This is also confirmed by the quite poor association of incidence gap with other classical measures of racial segregation (see Supplementary Figures 28-29).

The results presented in this work suggest that policy makers should definitely take into account mobility patterns when modelling the spread of a disease in a urban area, and in predicting the impact of specific countermeasures. In particular, a strategy to mitigate incidence gap should focus on reducing as much as possible longdistance trips for people that are naturally more exposed to face-to-face contacts, e.g., due to their occupation, and enforcing stricter measures of social distancing on local activities.

## METHODS

### Geographic network data sets

Ethnicity data was obtained from [34] and includes the data from the 2010 decennial census. Commuting trips data comes from the 2011 US census [35], focusing on the seven highest-level ethnicity classes, namely: White, Black or African American, American Indian and Alaska Native, Asian, Native Hawaiian and Other Pacific Islander, Some Other Race, Two or More Races. Population is updated to the latest American Community Survey 2014-2018 5-yeas Data Release [47].

For each metropolitan area we constructed two distinct spatial networks. The first one is the *adjacency network*, denoted by 𝒜 and obtained by associating each cell to a node and connecting two nodes with a link if the corresponding cells border each other. Notice that 𝒜 is an undirected and unweighted graph,. The second graph is the *commuting network*, denoted as 𝒞. In this network each node is a tract and the directed and weighted link *ω*_*ij*_ between node *i* and *j* indicates the number of commuting trips from *i* to *j* as obtained from census information. To reconstruct a mobility network that resembles the real one (which amounts to something between 30% and 40% of the total mobility in a city) we aggregated both the trips from home to work and the corresponding return trip from work to home.

Each node of the adjacency network 𝒜 preserves information about the ethnicity distribution on the corresponding census tract. We use the *N ×* Γ matrix *ℳ* = *{m*_*i,α*_ *}*, where Γ is the number of ethnicities present in the city. The generic element *m*_*i,α*_ of ℳ indicates the number of citizens of ethnicity *α* living on node *i*. We denote by *M*_*i*_ = *{m*_*i,α*_*}* the vector of population distribution at node *i*, and by 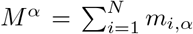 the total number of individuals of class *α* present in the system. In the commuting network, 𝒞 instead, we attribute to each node *i* both the resident population at the corresponding tract and the population commuting to node *i*, so that the abundance of individuals of class *α* on node *i* becomes:

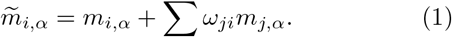

where *ω*_*ji*_ is the number of daily commuting trips from node *j* to node *i*. By doing so we aim to capture the fact that a commuter to cell *i* will potentially have face-to-face interactions with both residents in that area and other workers commuting to that area every day. Moreover, since the commuting network 𝒞 accounts for both work-home and home-work trips, the adjusted population on the commuting network accounts for the potential contacts that individuals had at the origin of a trip as well.

### Class Mean First Passage Time (CMFPT)

Let us consider a generic graph 𝒢 (𝒱, *ℰ*) with |𝒱| = *K* edges on |𝒱 |= *N* nodes, and a colouring function *f* : 𝒱 *→χ* that associates to each node *i* of 𝒢 a discrete label *f* (*i*) from the finite set *χ* with cardinality |*χ*| = Γ. Let us also consider a random walk on 𝒢, defined by the transition matrix Π = {*π*_*ij*_} where *π*_*ji*_ is the probability that the walk jumps from node *i* to node *j* in one step. On the adjacency 𝒜 network we use a uniform random walk, i.e., 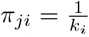, while on the commuting graph 𝒞 we have 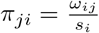, where *s*_*i*_ = ∑_*ij*_ *ω*_*ij*_ is the out-strength of node *i*.

Here we focus on the statistical properties of the trajectories *W*_*i*_ = {*f* (*i*_0_), *f* (*i*_1_), …} of node labels visited by the random walk *W* at each time when starting from *i*_0_ = *i* at time *t* = 0. This dynamics contains information about the existence of correlation and heterogeneity in the distribution of colours. For instance, if the graph *G* is a regular lattice and the function *f* associates colours to nodes uniformly at random, we expect that, for long-enough time, all the trajectories starting from each of the *N* nodes will be statistically indistinguishable.

We denote as *T*_*i,α*_ the Mean First Passage Time from a given node *i* to nodes of class *α*, i.e., the expected number of steps needed to a walk starting on *i* to visit for the first time any node *j* such that *f* (*j*) = *α*. We can write a self-consistent forward equation for *T*_*i,α*_ [48]:

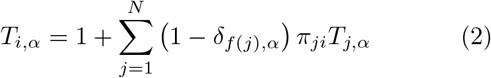

The Mean First Passage Time *τ*_*βα*_ from class *α* to class *β* is defined as:

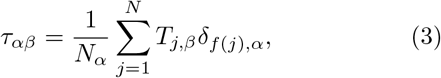

where *N*_*α*_ is the number of nodes in the graph associated to class *α*. Notice that in practice the value of 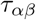 is obtained as an average over many realisations of the random walk.

A notable issue of the MFPT defined in Eq. (3) is the fact that its values might depend on the specific distribution of colours (i.e., on their abundance) and on the size of the network under consideration, which makes it difficult to compare Mean First Passage Times computed on different systems. To obviate to this problem, we define the normalised Class Mean First Passage Time between class *α* and class *β* as:

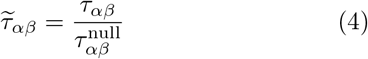

where 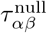 is the MFPT from class *α* to class *β* obtained in a null-model graph. The null-model considered here is the graph having the same topology of the original one, and where node colours have been reassigned uniformly at random, i.e., reshuffled by keeping their relative abundance. Notice that 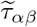 is a pure number: if 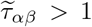 (resp., 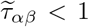 it means that the expected time to hit a node of class *β* when starting from a node of class *α* is higher (resp., lower) than in the corresponding nullmodel. In general, a value different from 1 indicates the presence of correlations and heterogeneity.

### Class Coverage Time (CCT)

The coverage time is classically defined as the number of steps needed to a random walk to visit a certain percentage of the nodes of a graph when starting from a given node *i* [48]. In the case of a network with coloured nodes, a walk started at node *i* will be associated to the generic trajectory 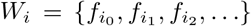 of node labels visited by the walk at each time. Since we are interested in quantifying the heterogeneity of ethnicity distributions, we consider the time series 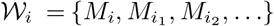 where 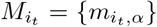 is the distribution of ethnicities at node *i*_*t*_ visited by the walk at time *t*. If we consider the trajectory up to time *t*, the vector 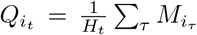 is the distribution of ethnicities visited up to time *t* by the walker started at *i* (here *H*_*t*_ is a normalisation constant that guarantees 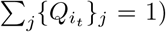. We quantify the discrepancy between 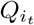 and the global ethnicity distribution across the city 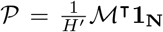 by means of the Jensen-Shannon divergence:

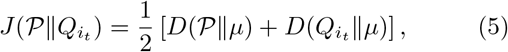

where 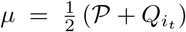 and *D*(*P* ∥*Q*) is the Kullback-Liebler divergence between *P* and *Q*. We define the Class Coverage Time from node *i* at threshold *ε* as:

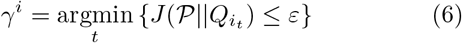

and the associated normalised Class Coverage Time:

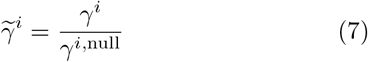

where *γ*^*i*,null^ is the Class Coverage Time from node *i* in a null-model where the colours associated to the nodes have been reshuffled uniformly at random.

### CMFPT and CCT in census networks

In the case of ethnicity distributions in geographical networks, each node is not uniquely associated to a colour, but it has instead a local distribution of ethnicities. Nevertheless, the formalism for the computation of Class Mean First Passage Times and Class Coverage Time described above can still be used in this case as well. We consider a stochastic colouring function 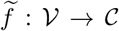 that associates to each node *i* of the adjacency graph one of the Γ = 7 ethnicities *α* with probability 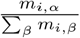 (respectively, with probability 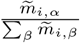; in the commuting graph), i.e., proportionally to the abundance of ethnicity *α* in node *i*.

To compute the CMFPT we consider *S* independent realisations of the stochastic colouring process for each network. On each realisation *ℓ*, we estimate the MFPT among all classes as in Eq. (3), and the corresponding null-model MFPT. Then, we compute the average Class Mean First Passage Time from class *α* to class *β* as:

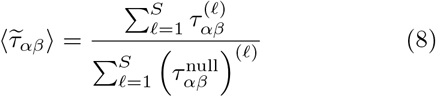

where 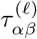 is the CMFPT computed on the *l*-th realisation and 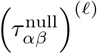 is the corresponding value in the null-model. For each system we computed 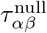 on 500 realisations of the null model, with 500 independent colour assignments per realisations, and 2000 walks per node.

The computation of CCT works in a similar way. In order to take into account the heterogeneous distribution of ethnicities across nodes, before a walker starts from node *i* we sample one of the ethnicities present on *i*, according to their local abundance at *i{m*_*i,β*_ *}*, and we attribute node *i* to it. Then, we compute the CCT from node *i* of class *α* as the average CCT from node *i* across all the walks starting from *i* where node *i* was actually assigned to class *α*, and we call this quantity 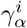. Notice that in this case we consider the trajectories 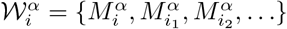 where 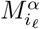 is the distribution of ethnicities at the *ℓ*-th node visited by the walker, which does not include class *α*. The normalised Class Coverage Time from class *α* when starting from node *i* is defined as:

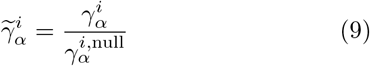

where 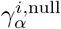 is the CCT from node *i* of class *α* in the nullmodel. Finally, the average CCT from class *α* is simply
obtained as:

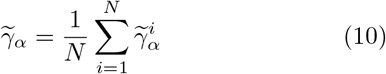

For all the computations of CCT shown in the paper we considered averages over 5000 walks per node and we set *ε* = 0.0018. This value was obtained by using the Pinsker’s inequality for the Kullback-Leibler divergence and imposing a total variation distance smaller than 6%.

### Global indices of dynamic ethnic segregation

We constructed three global indices of dynamic segregation based on the values of CMFPT and CCT. In particular, we focused on the observed discrepancies of CMFPT and CCT between African Americans and other ethnicities. In the following the index *A* will always indicate African Americans, while the index *O* will indicate all the other ethnicities. We start by defining the following quantities:

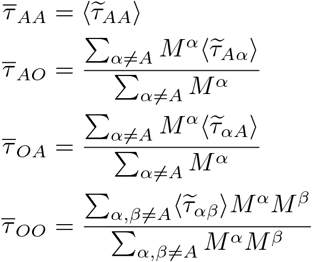

In practice: 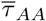 is the expected CMFPT from African Americans to African Americans; 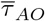 is the expected CMFPT from African Americans to all the other classes (weighted by ethnicity distribution); 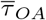 is the expected CMFPT from all the other classes to African Americans (again, weighted by ethnicity distribution); and 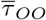 is the expected CMFPT among all the other ethnicities. Notice that all these quantities are pure numbers, since they are based on the corresponding quantities defined in Eq. (8) which are correctly normalised with respect to the null-model.

The clustering of African Americans is quantified as:

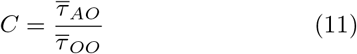

so that values of *C* larger than 1 indicate that for an African American finding any other ethnicity is harder (i.e., requires more time) than for all other ethnicities. Similarly, we define the exposure of African Americans to other ethnicities as:

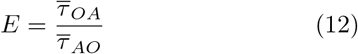

where values of *E* larger than 1 indicate that it is easier for African American to be found in touch with any other ethnicity than for people from all the other ethnicities to be found in touch with African Americans.

We define similar quantities for the *CCT* of African Americans and other ethnicities, namely 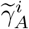 as in Eq. (9), and:

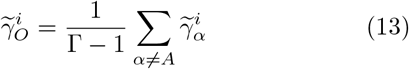

Finally, we define the isolation of African Americans for the whole system by the average ratio:

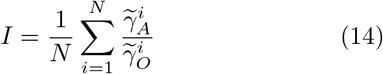

over all the nodes. Notice that values of *I* larger than 1 indicate that the normalised CCT from nodes of class *A* (African American) is higher than the CCT from nodes of all the other classes. The State-level value of each index is obtained as an average of the corresponding index on the cities of the state, weighted by the population of each city.

### Local dynamic ethnic segregation

We define two local segregation indices for African Americans in a census tract *i*. The first index is based on CMFPT:

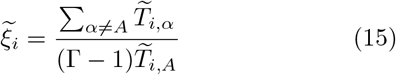

where 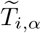 corresponds to the normalised Mean First Passage Time to a generic class *α* when a random walker starts from node *i*, while 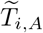 is the CMFPT to African Americans tracts. Values of 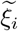 larger than 1 indicate that the time to reach any other ethnicity is higher than the time needed to reach African Americans, hence indicating a local clustering of African Americans around node *i*.

The local index of isolation is derived from CCT:

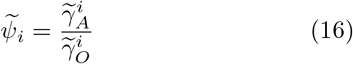

where 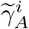 is the CCT from node *i* for African Americans and 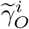 is the average CCT from node *i* for all the other ethnicities, as defined in Eq. (13). In general, if 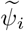 is larger than 1 then African Americans living at node *i* are isolated, since they will require more time to visit all the other classes than required by individuals from other ethnicities.

### Data on COVID-19 incidence among the African American Population

The data related to the percentage of infected African Americans was obtained from two different sources [32] and [33]. The first data set reports the number of infected and deceased of each ethnicity along with those unknown. To calculate the percentage of African Americans we have removed first the unknown from the total, otherwise our analysis would also capture the fraction of unknown. For the other data set we just extract the data they provide in tables.

### Public Transportation data set

The public transportation data set was obtained from the 2018 American Community Survey from U.S. Census Bureau [34]. It includes information about the percentage of public transportation usage per ethnicity and State.

### Multivariate Analysis

The multivariate analysis was performed using R and the ANOVA model in the car package.

## Data Availability

All the data used in this work was obtained from public sources. No new data sets weregenerated in this study. The processed data sets are available from the corresponding author on reasonable request.

## ACKNOWLEDGEMENTS

A.B. and V.N. acknowledge support from the EPSRC New Investigator Award Grant No. EP/S027920/1. This work made use of the MidPLUS cluster, EPSRC Grant No. EP/K000128/1. This research utilised Queen Mary’s Apocrita HPC facility, supported by QMUL ResearchIT. doi.org/10.5281/zenodo.438045.

## AUTHOR CONTRIBUTIONS

All the authors devised the study. A.B. and S. S. performed the simulations and computations. All the authors provided methods and analysed the results. A. B. and S. S. prepared the figures and all the visual material. All the authors wrote the paper and approved the final submitted version.

## Appendix A Quantifying ethnic segregation through CMFPT and CCT

### Ethnic segregation and COVID-19 incidence through CMFPT

Ethnic segregation is quantified here through random walks, and more precisely, class mean first passage times (CMFPT). Given a set of classes present in a city – ethnicities in this case– we are interested in the number of steps you need to reach one as a function of the ethnicity at the origin. Random walks start from each of the city cells –or tracts– and move until they have visited each of the distinct classes or ethnicities present in a city. If we average the passage times across all the city cells and then divide by the same quantity from the null model we obtain the normalised CMFPT between class *α* and 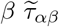. It is important to note that this matrix is not necessarily symmetric and depends on the spatial distribution of classes. We show in Supplementary Figure S-1 the CMFPT on the adjacency graph for four cities: Detroit, Chicago, Houston and Los Angeles. On a first look, strong differences can be detected between those cities on the left and those on the right. The normalised CMFPT are substantially higher in Detroit and Chicago when compared to Houston and Los Angeles. Despite the difference in the maximum values, the shape of the matrix and curves is not so different across cities, with African Americans much more isolated than the rest of ethnicities. Reaching African Americans is much harder for any other class, while it is considerably low for other African Americans. As can be seen, there are not only strong differences between both cities but also between the type of network used (See Supplementary Figure S-2). One significant change that appears in some cities when 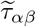 is computed over the adjacency graph is that for African Americans, Whites are more easy to reach than themselves, which means that mobility plays a crucial role on approaching African Americans to the rest of the population and exposing them. Additionally, the normalised CMFPT seems to be less dependent on the ethnicity of the origin and more on the ethnicity of the destination. Likely as a consequence of the higher mixing produced by the long-range links present in the mobility network. Not only that, but the differences between each ethnicity are also reduced. Overall, to properly quantify segregation we need to take into account not only the residences but also how the ethnicities move in cities.

### Ethnic segregation and COVID-19 incidence through CCT

We consider a number of Consolidated Statistical Areas (CSA) in the US, 128 networks are constructed based on adjacency while 171 networks were constructed for commuting. These systems are represented as a spatial graph 𝒢 and we look at the statistical properties of the trajectories of a random walk on 𝒢. Each walk starting at node *i* is associated to an ethnicity sampled from 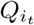 and it stops at time *t* when 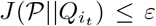. When the ethnicity *α* is sampled, the corresponding bin is removed from the computation 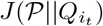 so that the effect that *α* has on the coverage time at threshold *E* can be quantified.

**FIG. S-1.**
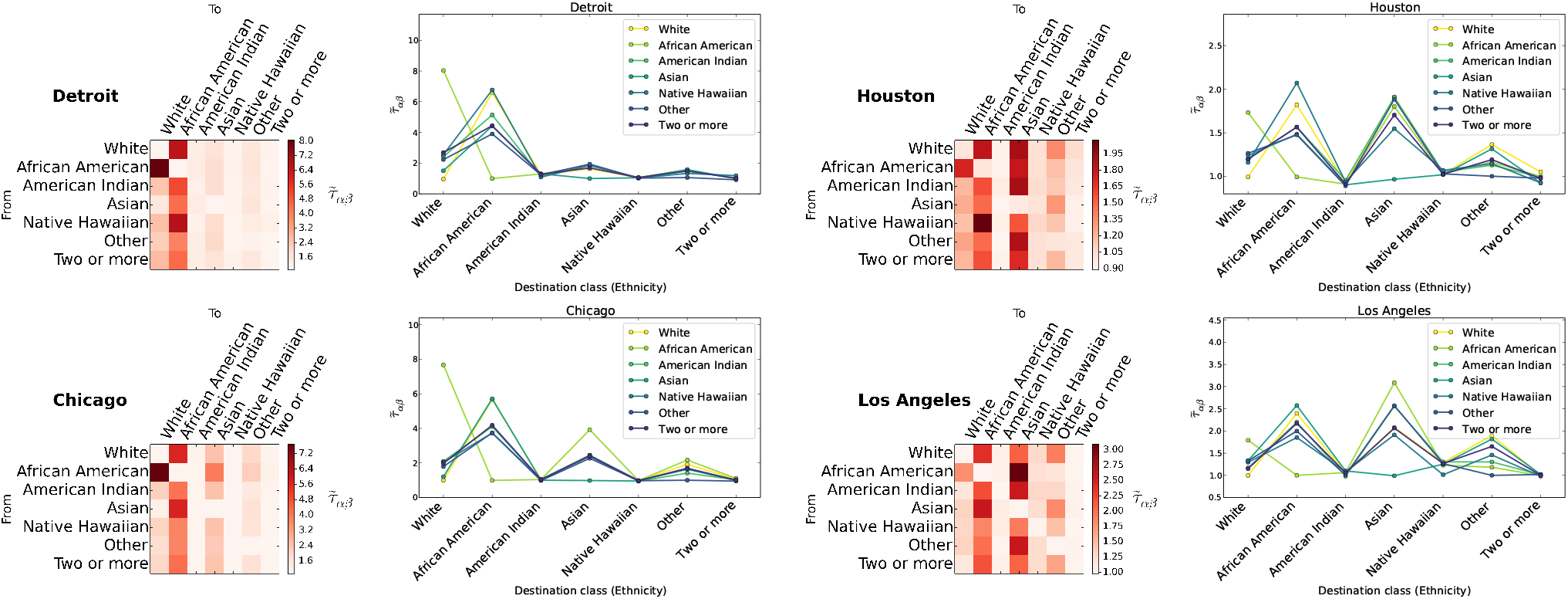
Normalised inter-class mean first passage times among the different ethnicities contained in our data set when walkers move on the adjacency network in two different visualisation styles for the following cities: Chicago, Detroit, Houston and Los Angeles.

**FIG. S-2.**
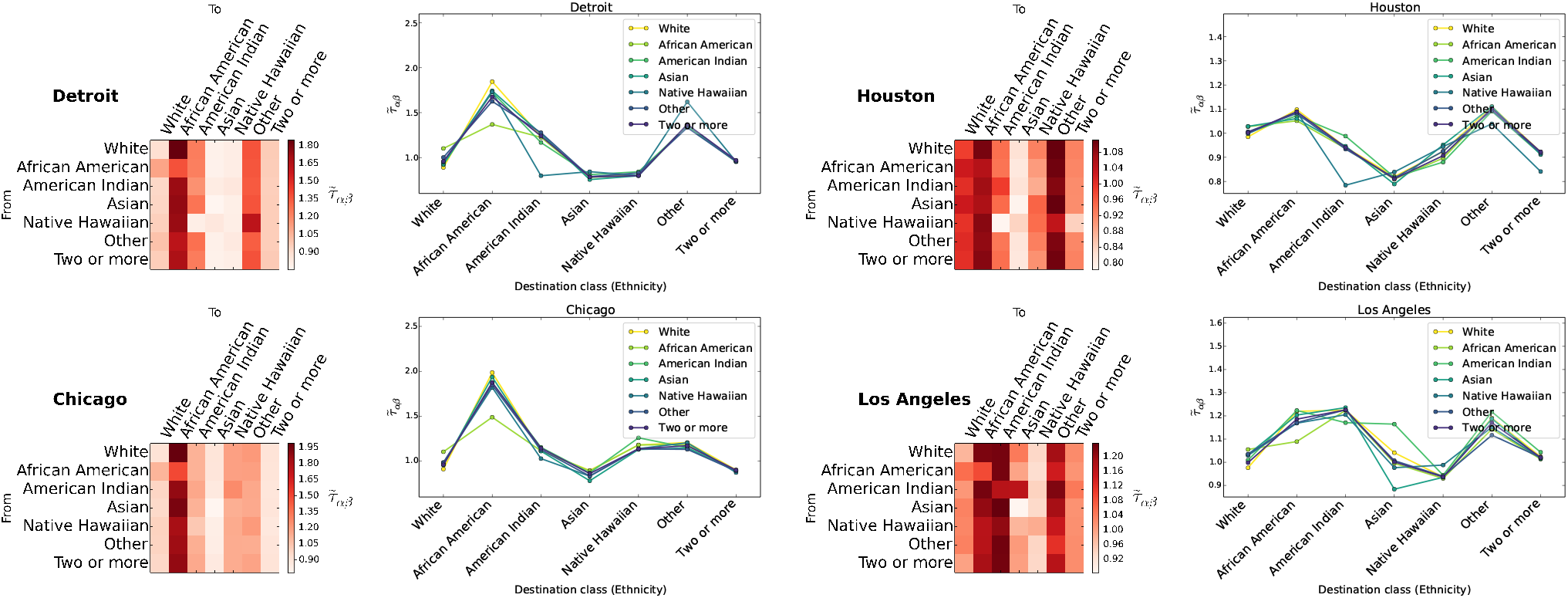
Normalised inter-class mean first passage times among the different ethnicities contained in our data set when walkers move on the commuting network in two different visualisation styles for the following cities: Chicago, Detroit, Houston and Los Angeles.

Trajectories from each node are averaged over 5000 repetitions for the adjacency network and 2000 for commuting. The null model for *CCT* is obtained by randomly reassigning the vector *M*_*i*_ to a new node in and 𝒢 the concentration of an ethnicity in a region if such spatial pattern is present is dissolved across 𝒢. Here we consider 20 independent repetitions of the null model for each CSA on both adjacency and commuting networks, then, the Class Coverage Time (*CCT*) for the null model of a city is the average behaviour over 20 independent realisations.

The normalised *CCT* is reported for the two spatial configurations in Fig. S-3 **a-b**. The coverage times obtained for the adjacency networks **a** are considerably larger compared to commuting **b**, where on the later, the majority of values spams in a small range between 0 and 4. The distributions of 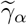 for each ethnicity have a comparable shape within the network type where most values are contained in a common interval, yet, they are distinguishable and differences between the ethnicities can be observed. The corresponding non-normalised quantities can be read on panels **c-d** where the *CCT* of the real system and the equivalent null model are reported.

**FIG. S-3.**
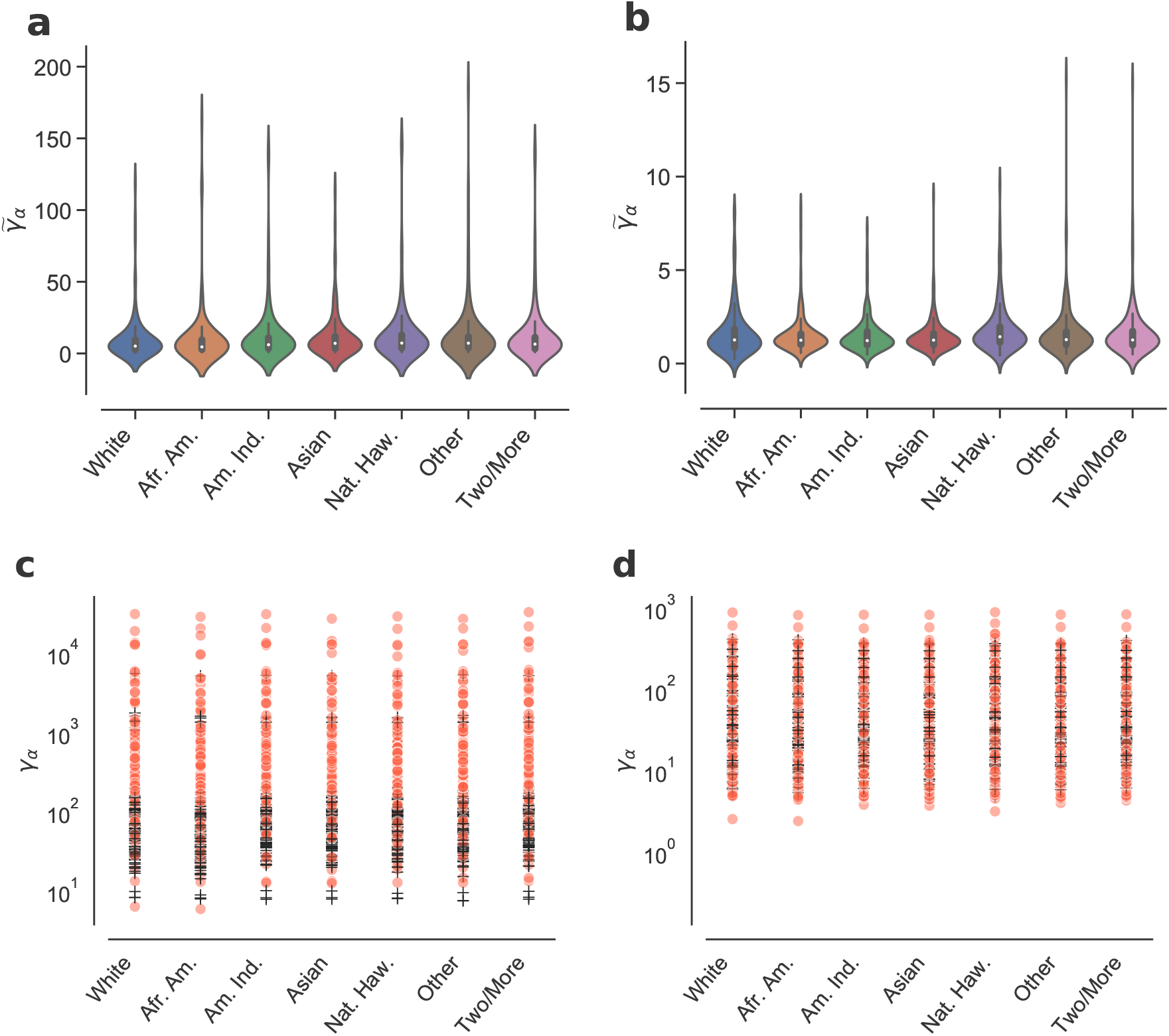
Class coverage times and the corresponding normalised values on the adjacency **a** and commute **b** networks for all CSA. Panels **c-d** report the coverage times (non-normalised) for adjacency and commute networks where the black crosses correspond to the values for the equivalent null model. Each data point contained in an ethnicity column is equivalent to 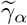 at a CSA.

It is important to note that the difference on the *CCT* of the real system and the null model is significantly large on the adjacency networks, with the former having cases where coverage times are two orders of magnitude larger (See Fig. S-3 **c**). Although the null model corresponds to the non-segregated counterpart of the city and coverage times are expected to be smaller, these large differences suggest caution and open an interesting question for further investigation. In particular, to understand what factors influence the large differences, for instance if it is mainly driven by the population distribution, the threshold *ϵ*, the network topology or the combination of two or more factors.

In addition to the cities discussed in the main manuscript, we report two other systems in Fig. S-4 where individual values of 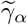 for each ethnicity can be observed on the adjacency and commute networks. African Americans are substantially less isolated in Houston compared to the other ethnicities in both adjacency and commute networks. In Detroit, The adjacency information gives the opposite picture for African Americas while Whites are the most isolated on the commute network.

**FIG. S-4.**
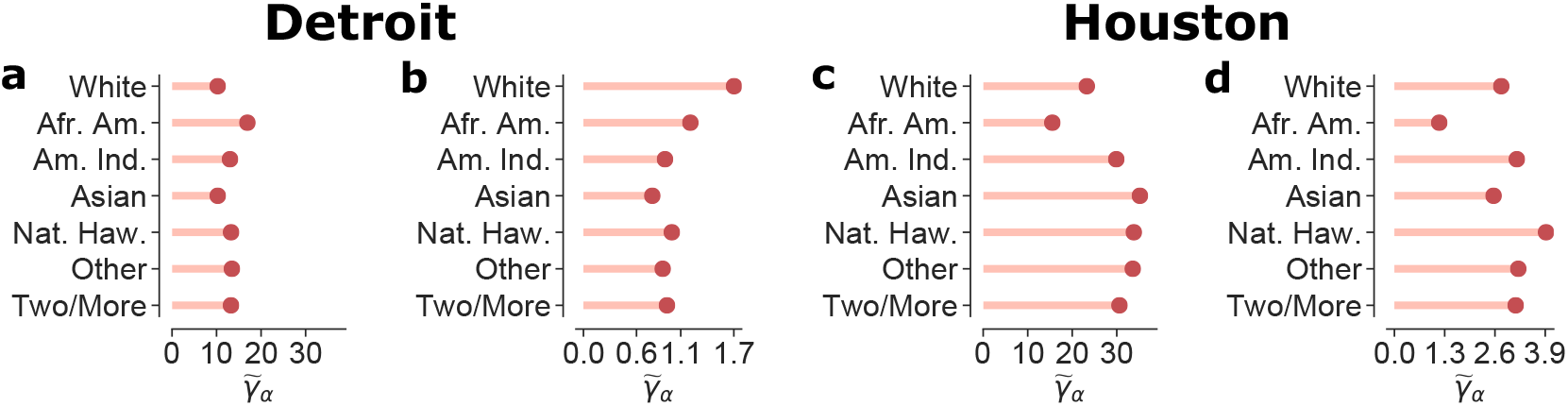
Normalised coverage time on the adjacency and commute networks for Detroit **a-b** and Houston **c-d**. Values are larger on the adjacency network compared to commute for both cities while African Americans are significantly less isolated in Houston for both networks.

**TABLE S-I.**
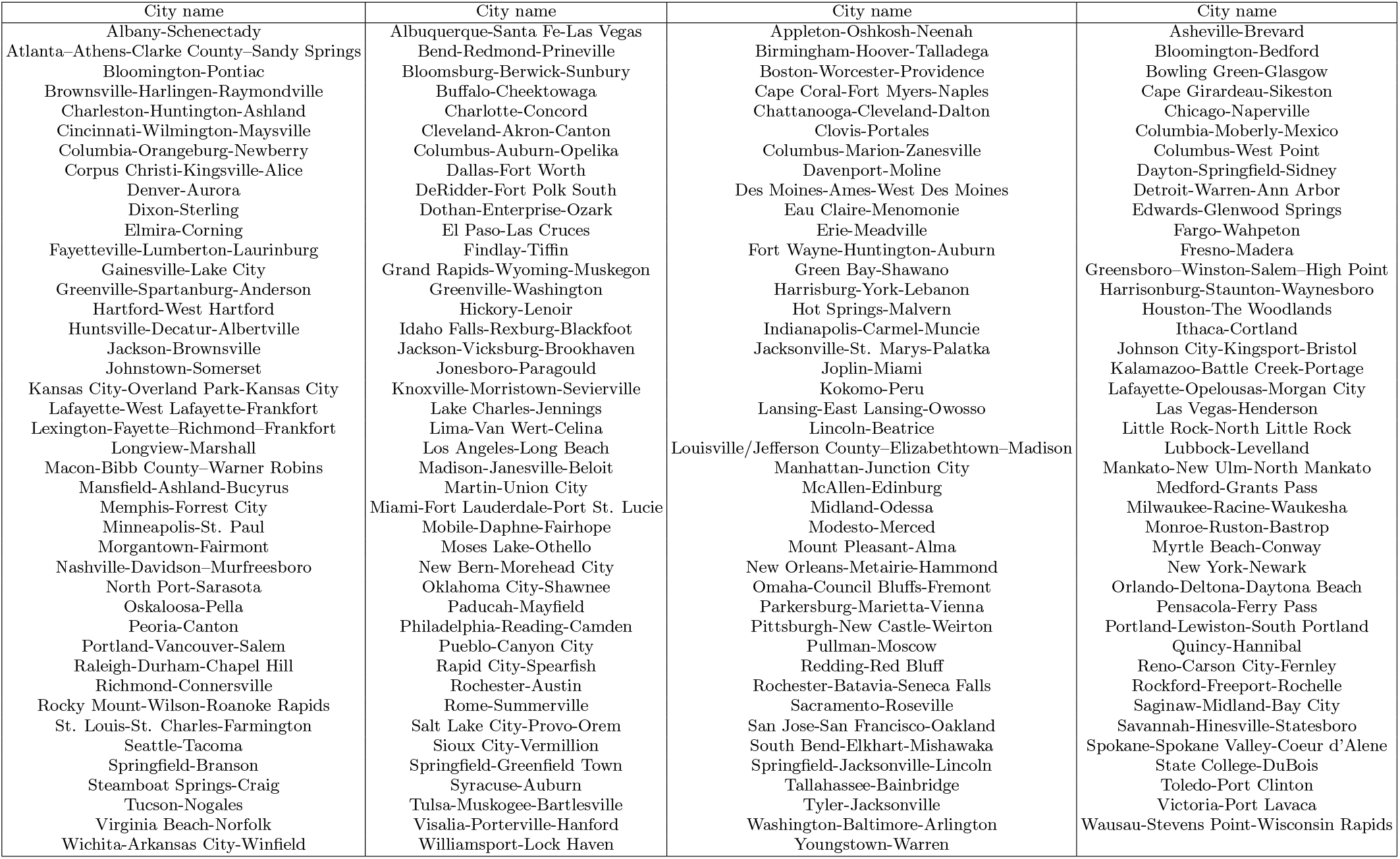
Table of cities studied

## Appendix B Correlations between the incidence of COVID-19 among the African American population cases and ethnic segregation)

### Rankings and comparisons

We detail the cities studied in Supplementary Table S-I, it is important to note that those are the cities studied disregarding if those states provide ethnic information on the impact of COVID-19.

Supplementary Figure S-5 displays the ranking of values for each of the four metrics studied in the main manuscript computed over the adjacency or commuting graphs. As can be seen, strong similarities between rankings appear.

To evaluate how similar are those rankings we have calculated the Kendall *τ*_*k*_ between each pair of rankings. Supplementary Figure S-6 displays the values of *τ*_*k*_ between each pair of the four metrics studied in the main manuscript computed in the adjacency and commuting graphs. For instance, there is a high correlation between the index *C* computed in the adjacency and the commuting graphs while for the exposure index *E* there is almost no correlation between both. Likely pointing out that exposure can only be effectively measured by including the commuting network. Another additional observation is the connection between *C* and *E* measured on the commuting graph, which seems to point out that in those states where African Americans are more segregated they are also more exposed.

**FIG. S-5.**
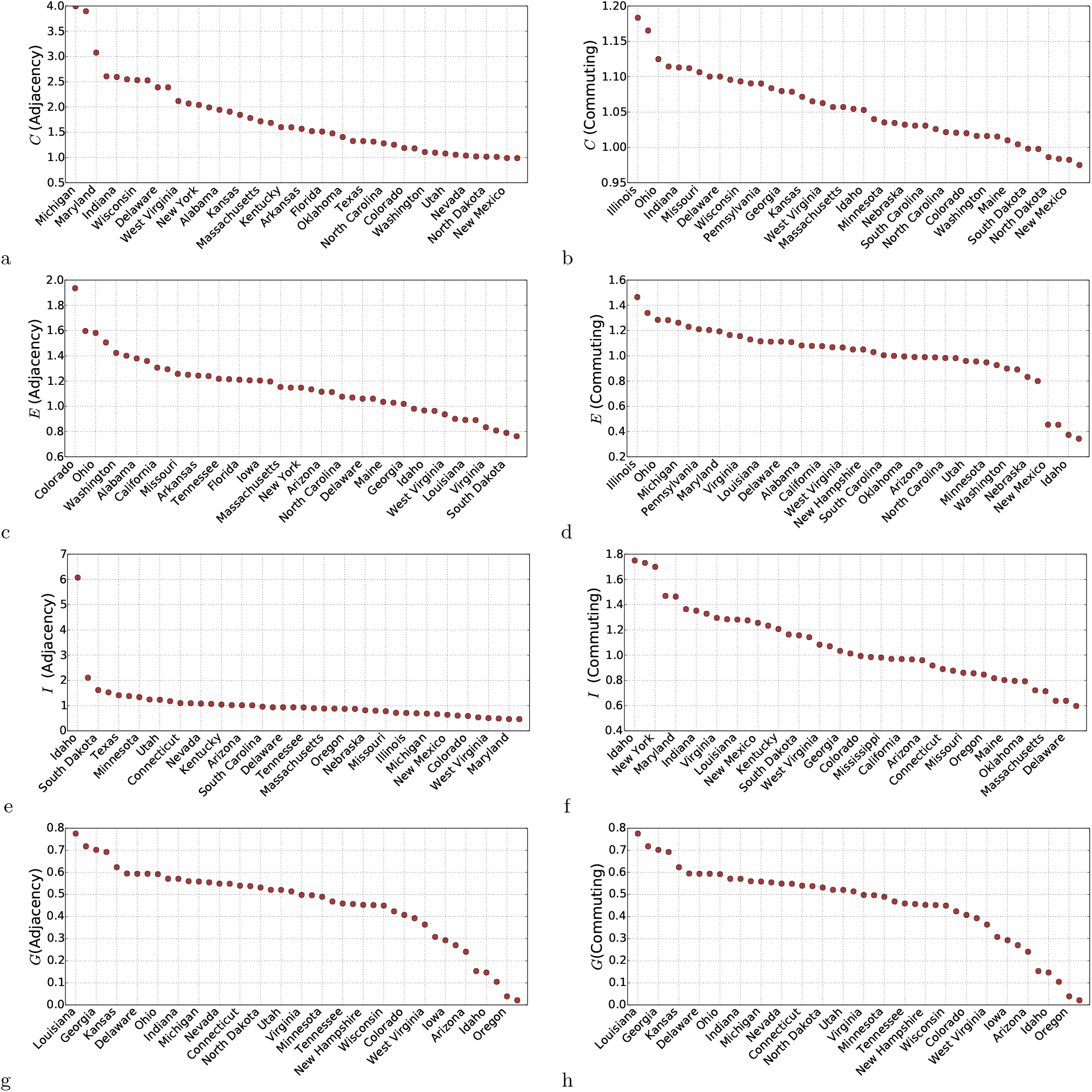
Ranking for the four indices studied in the main manuscript: *C, E, I* and*G* computed in both the adjacency and commuting graph. **a** *C* (Adjacency), **b** *C* (Commuting), **c** *E* (Adjacency), **d** *E* (Commuting), **e** *I* (Adjacency), **f** *I* (Commuting), **g** *G* (Adjacency), **h** *G* (Commuting).

In Supplementary Figure S-7, we compare the indices studied in this work, showing that most of them are related to each other yet not necessarily linearly. We find that those indices capturing the clustering of African Americans are also related to those related to mixing and exposure. The more clustered together, more sensible and exposed to the rest of the population. When comparing the same indices in the commuting and adjacency graphs, we observe that they have a non-linear relation, highlight again the importance of considering both of them.

**FIG. S-6.**
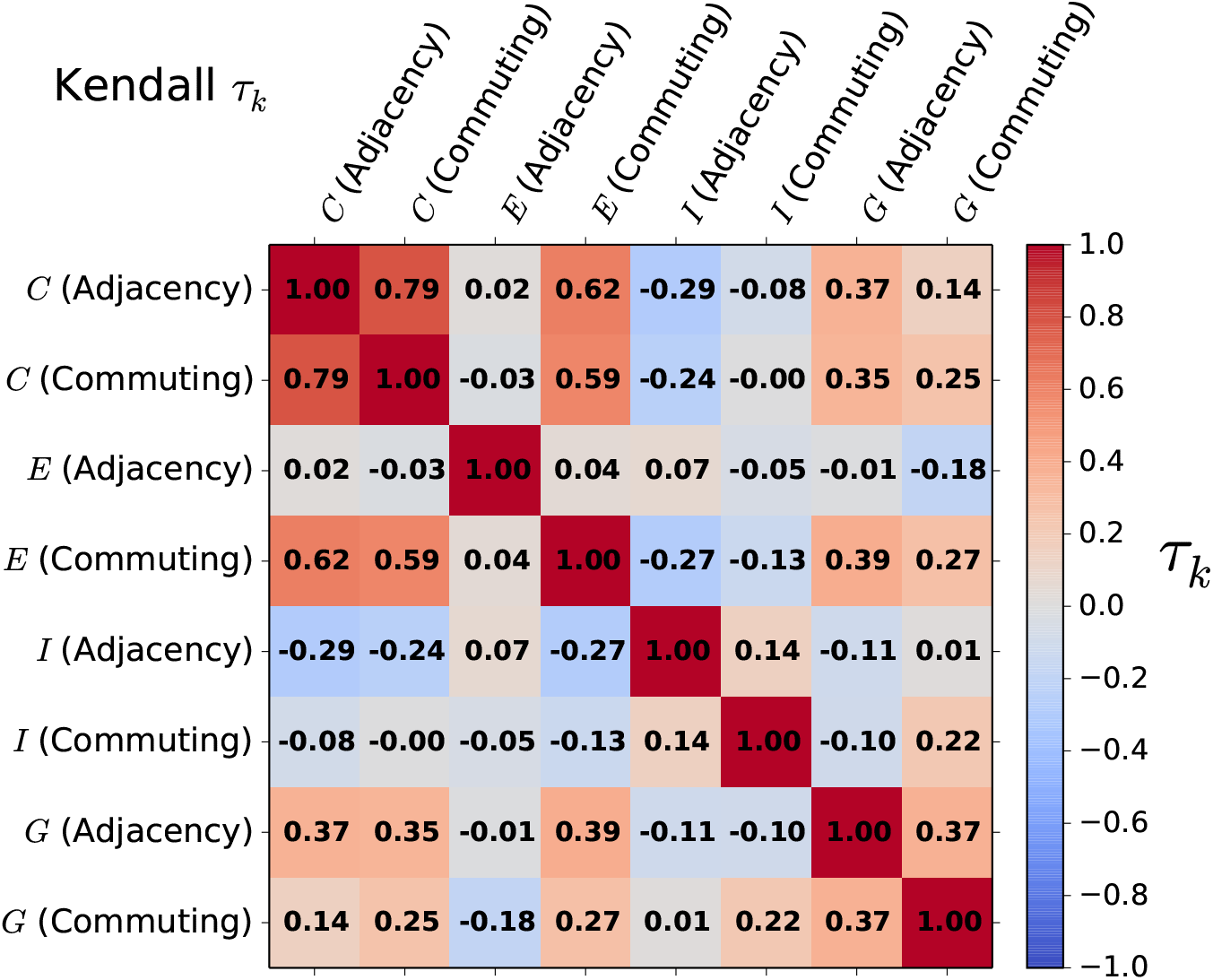
Kendall tau *τ*_*k*_ correlation between each of the four indices studied in the main manuscript computed either over the adjacency or the commuting graphs.

### Correlations between the segregation indices and other measures of COVID-19 incidence

The COVID-19 data used was obtained from [32] and includes several temporal snapshots until mid-may. The main variables we used are the difference on infected/deceased African Americans, where 0 would mean that the percentage of African Americans in the population of a state is the same than the percentage of infected, and the ratio calculated as the percentage of infected African Americans divided by their percentage among the overall population, which will be one if they are equal and higher than one if there are more African Americans infected/deceased among the overall population. Supplementary Table S-II summarises the results obtained for the linear fit for the Figure 2 in the main manuscript.

Supplementary Figure S-8 summarises the results obtained in the case of the ratio of infected African Americans. As detailed in the main manuscript, there are two versions of each index depending on whether the walkers move upon the adjacency or the commuting network. Compared to the difference in percentage correlations are much lower for the ratio, likely as a consequence of the several outliers. While states with a low percentage of African Americans among the overall population might easily suffer a huge increase on the ratio, those with a higher percentage of African Americans among the population might have a lower increase.

We have also evaluated how our metrics relate to the ratio and difference among the deceased African Americans (See Supplementary Figures S-9 and S-10). Despite many more factors such as the age or underlying health conditions might influence the deceased individuals, still, most of the correlations remain significant to some extent, especially those related to their exposure. Moreover, those indices computed on the commuting network seem to be more informative than those based on the adjacency, which seems to point out that residential segregation provides only a partial picture of ethnic inequality. Mobility is also crucial to understand the mixing between different ethnicities, it is not only relevant where certain ethnicities live but also where they work and with whom they interact when they do so.

Overall, despite our metrics are informative in both cases, they seem to be more related to the difference in percentage more than the ratio. There are states in which the percentage of African Americans among the population is low and, therefore, the ratio can increase drastically.

**FIG. S-7.**
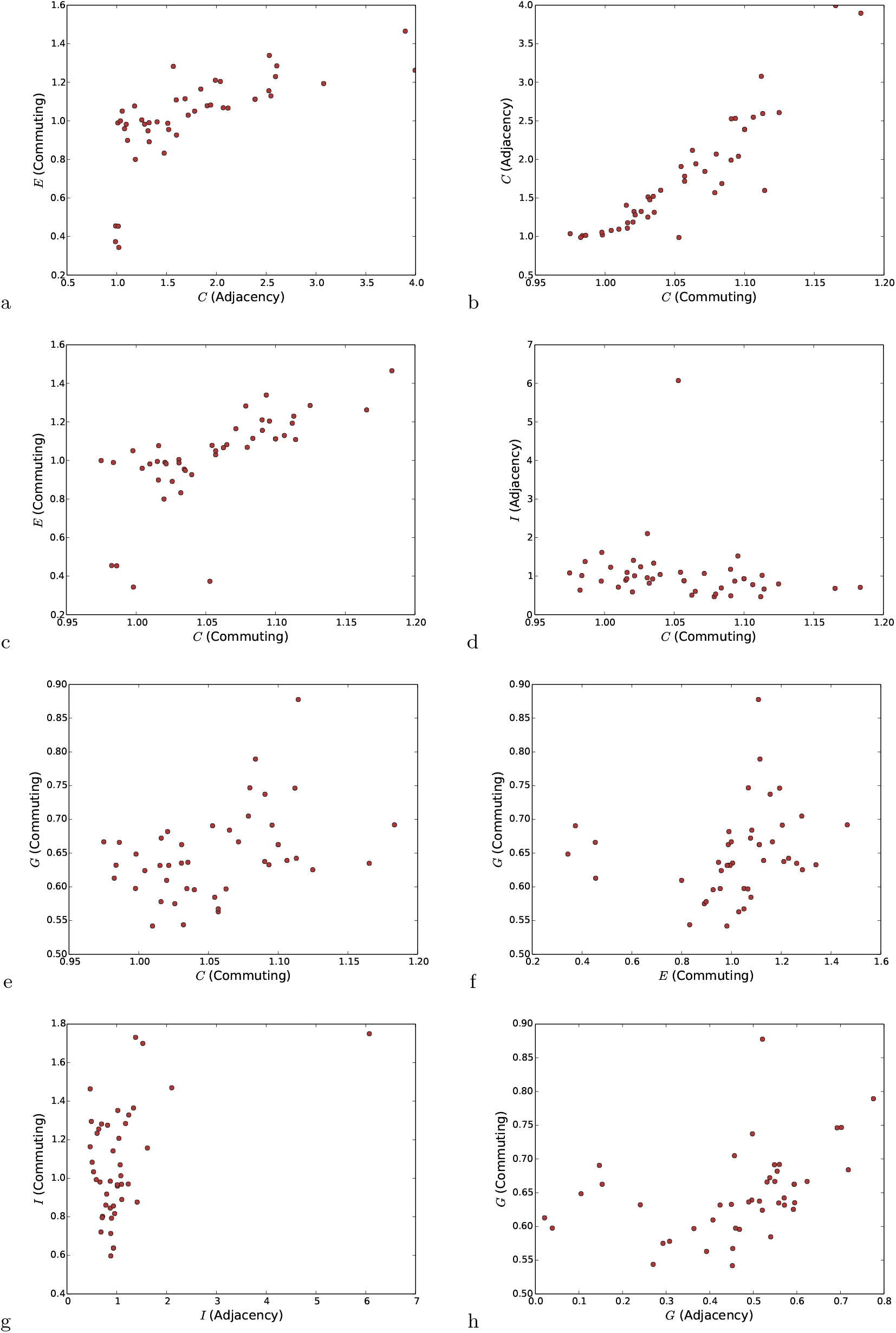
Comparison between the indices studied in the main manuscript. **a** *C* (Adjacency) and *E* (Commuting), **b** *C* (Commuting) and *C* (Adjacency), **c** *C* (Commuting) and *E* (Commuting), **d** *C* (Commuting) and *I* (Adjacency), **e** *C* (Commuting) and *G* (Commuting), **f** *E* (Commuting) and *G* (Commuting), **g** *I* (Adjacency) and *I* (Commuting), **h** *G* (Adjacency) and *G* (Commuting)

**FIG. S-8.**
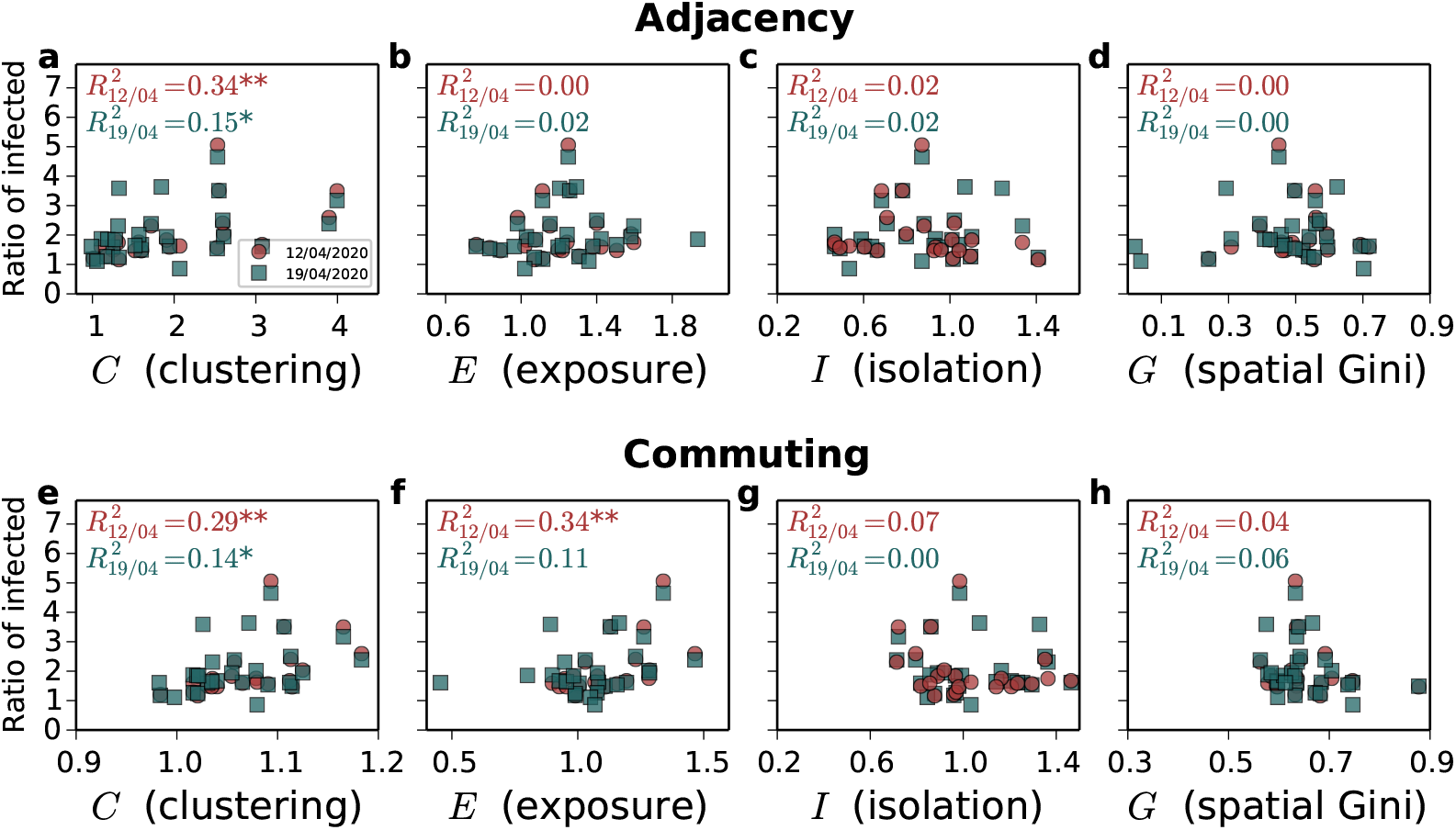
**Relation between the ratio of infected African Americans and the four indices considered. a-g** Indices computed over the adjacency network: **a** *C* (clustering), **b** *E* (exposure), **c** *I* (isolation), **d** *G* (spatial Gini). **e-h** Indices computed over the commuting network: **e** *C* (clustering), **f** *E* (exposure), **g** *I* (isolation), **h** *G* (spatial Gini). Each of the colours corresponds to a temporal snapshot of the data set, red for 12*/*04*/*2020 and blue for 19*/*04*/*2020. The *R*^2^ is computed as the square of the linear correlation coefficient.

**FIG. S-9.**
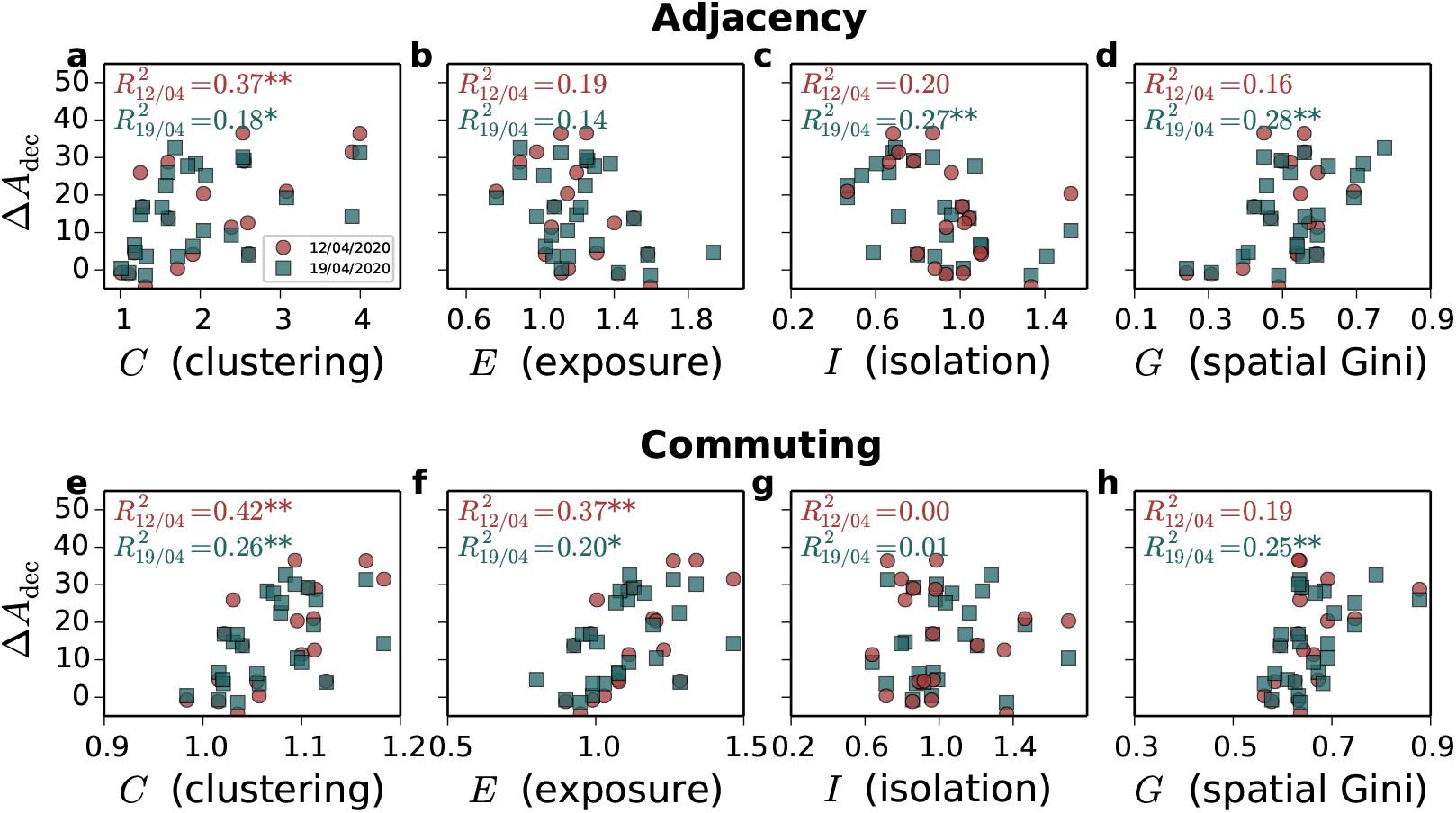
**Relation between the difference on the percentage of deceased African American as a function of the four indices considered. a-g** Indices computed over the adjacency network: **a** *C* (clustering), **b** *E* (exposure), **c** *I* (isolation), **d** *G* (spatial Gini). **e-h** Indices computed over the commuting network: **e** *C* (clustering), **f** *E* (exposure), **g** *I* (isolation), **h** *G* (spatial Gini). Each of the colours corresponds to a temporal snapshot of the data set, red for 12*/*04*/*2020 and blue for 19*/*04*/*2020. The *R*^2^ is computed as the square of the linear correlation coefficient.

**TABLE S-II.**
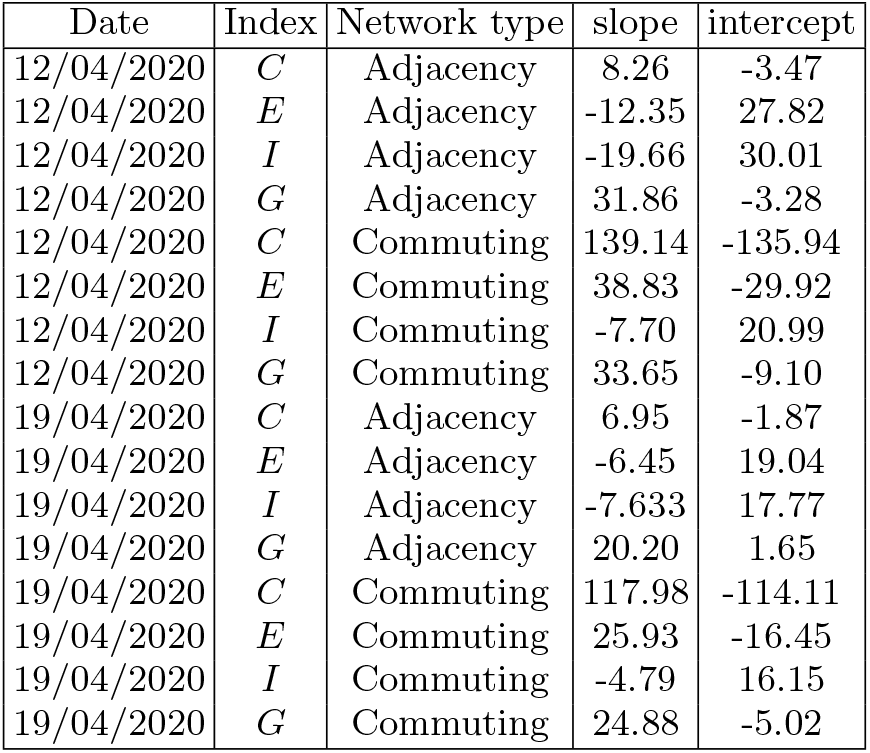
Coefficients obtained from the linear fits in Figure 2 of the main manuscript

**FIG. S-10.**
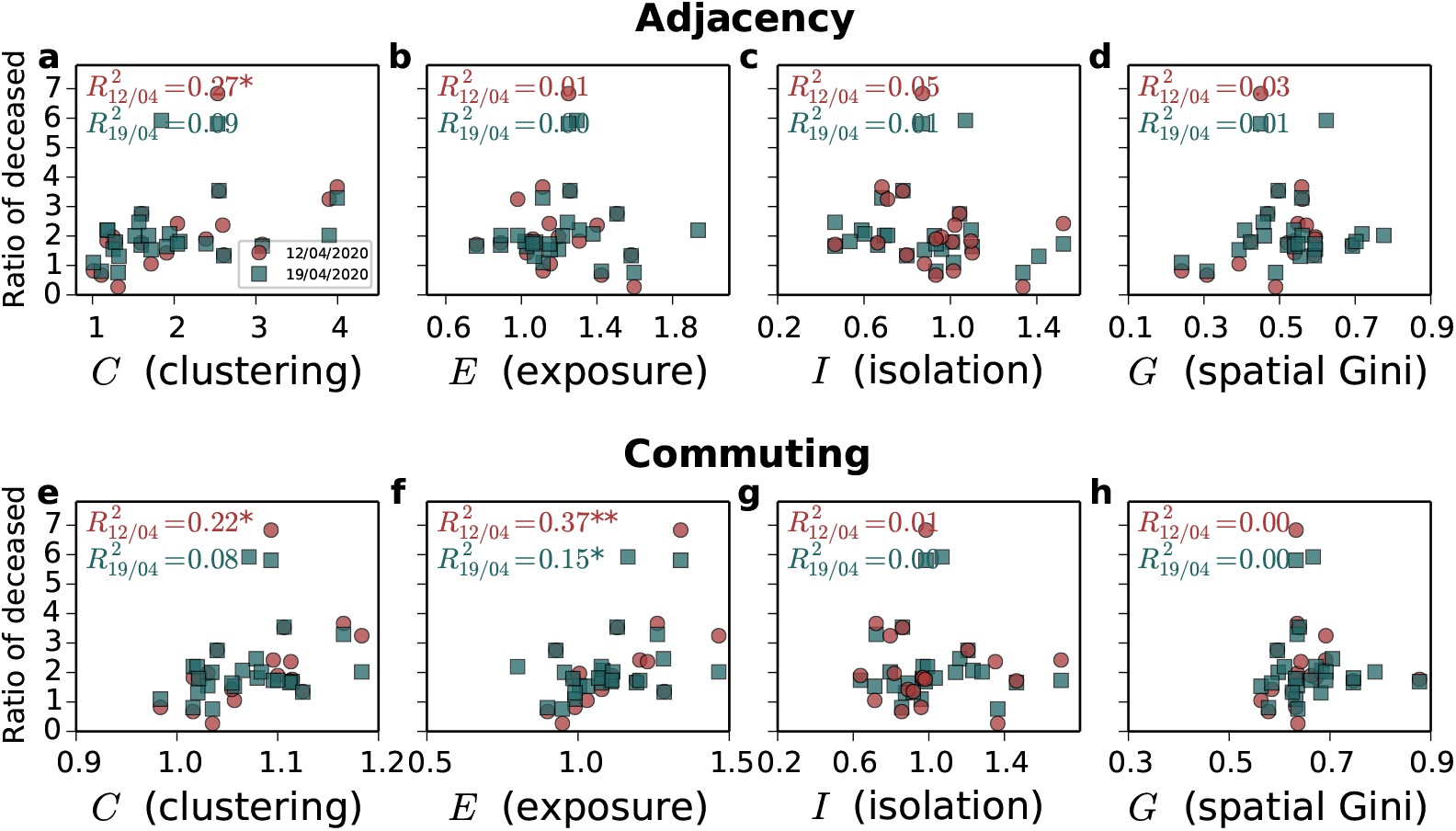
**Relation between the ratio of deceased African Americans and the four indices considered. a-g** Indices computed over the adjacency network: **a** *C* (clustering), **b** *E* (exposure), **c** *I* (isolation), **d** *G* (spatial Gini). **e-h** Indices computed over the commuting network: **e** *C* (clustering), **f** *E* (exposure), **g** *I* (isolation), **h** *G* (spatial Gini). Each of the colours corresponds to a temporal snapshot of the data set, red for 12*/*04*/*2020 and blue for 19*/*04*/*2020. The *R*^2^ is computed as the square of the linear correlation coefficient.

Ideally, each ethnicity *α* should be compared with the corresponding ratio to the overall population and the incidence of COVID-19 cases. As this data was not available during the preparation of this work, we look at the gap Δ*A*_inf_ of African American and the relation with the Isolation level of all other ethnicities in this study. Considering the quantity for all other ethnicities defined as:

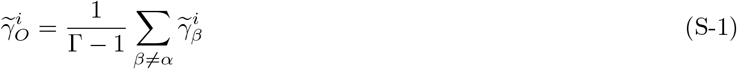

the Isolation index for an ethnicity *α* is given by:

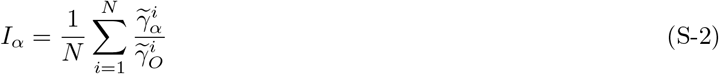

Correlations with the infection rate gap Δ*A*_inf_ on the adjacency and commute network are reported on Fig. S-11 and S-12 for all ethnicities. Quantities are obtained from the COVID-19 data set at 2 different periods, 12-04-2020 and 19-04-2020 respectively. The corresponding *R*^2^ of the Pearson correlation is reported in the inset of each panel. There is a negative correlation in **b** which indicates that less isolated African Americans have a higher incidence of infection cases. Whites **a** and Native Hawaiians **e** exhibit no correlation while the remaining ethnicities **c-d** and **f-g** have a positive *R*^2^ which decreases over time. We found no correlation for any ethnicity on the commuting network.

**FIG. S-11.**
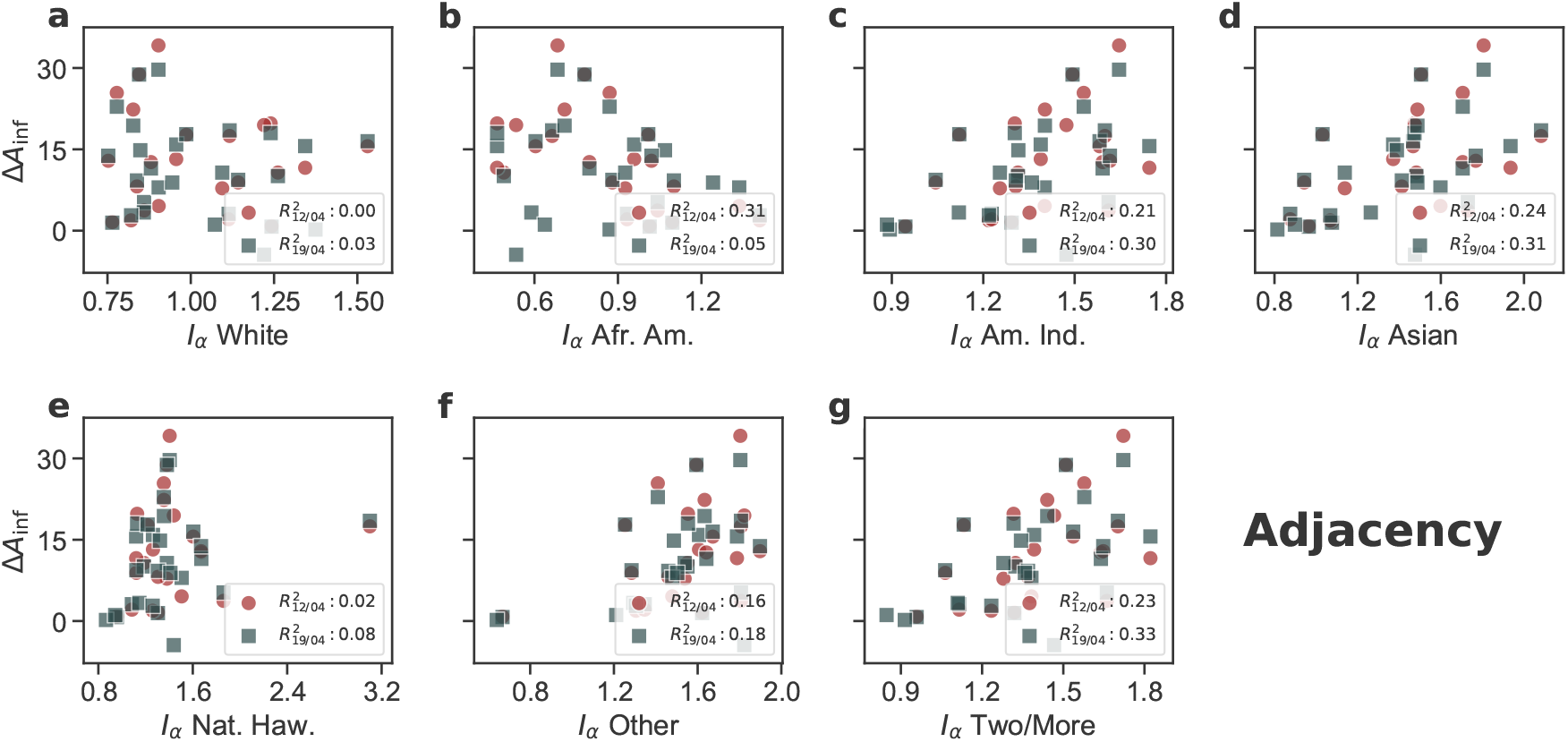
Isolation index of all ethnicities as a function of the infected rate gap of COVID-19 cases in the African American population on the adjacency network. African American is the only ethnicity to exhibit a negative correlation of isolation and Δ*A*_inf_, suggesting that a higher infection rate can be related to lower isolation.

**FIG. S-12.**
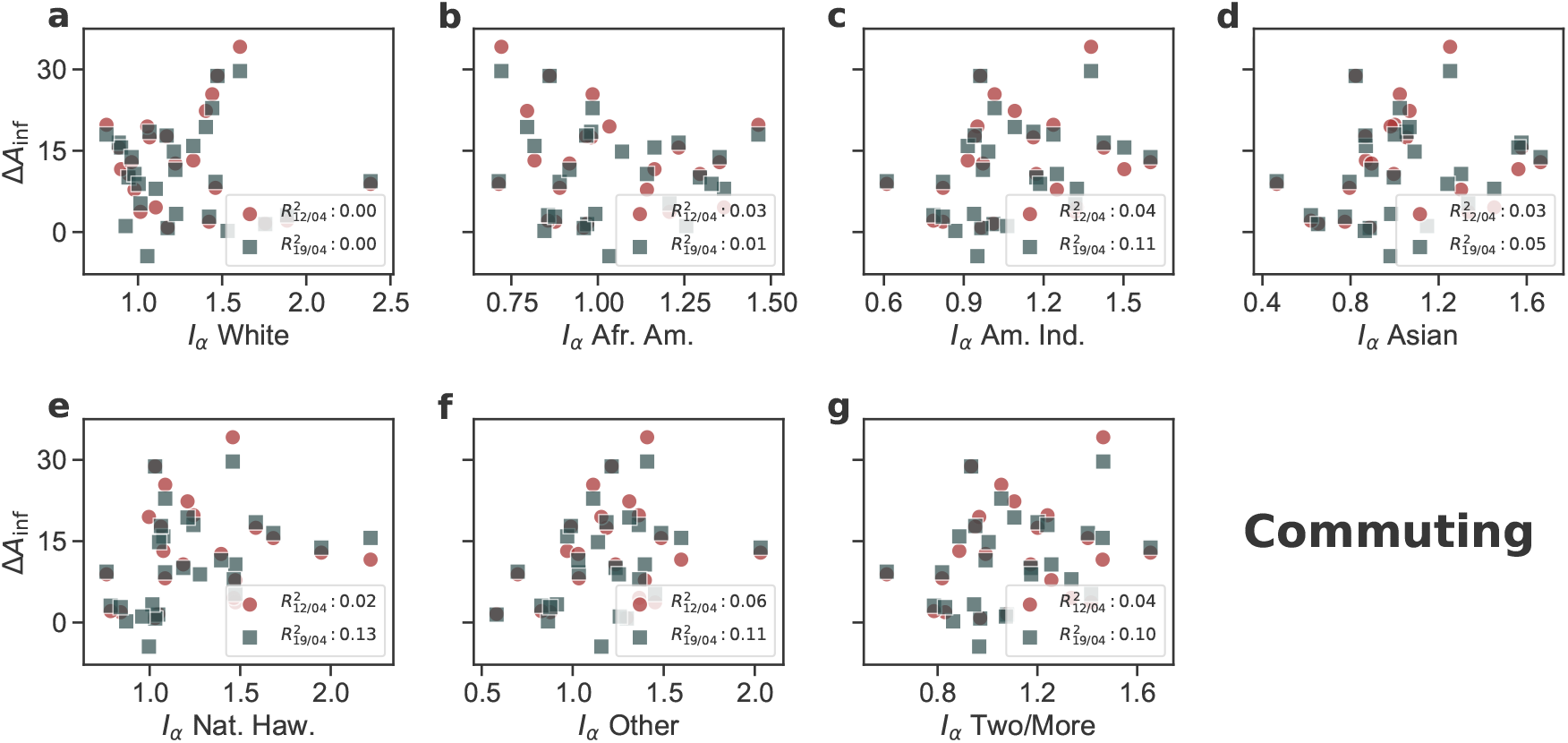
Isolation index of all ethnicities as a function of the infected rate gap of COVID-19 cases in the African American population considering the commute network. There is no significant correlation for any of the ethnicities.

Similarly, correlations with Δ*A*_deceased_ are computed for the deceased data on the adjacency and commute networks (See Fig. S-13 and S-14). We can observe a similar pattern with the results obtained from infected rates where there is correlations for the same group of ethnicities and no significant relationship on the commuting network.

**FIG. S-13.**
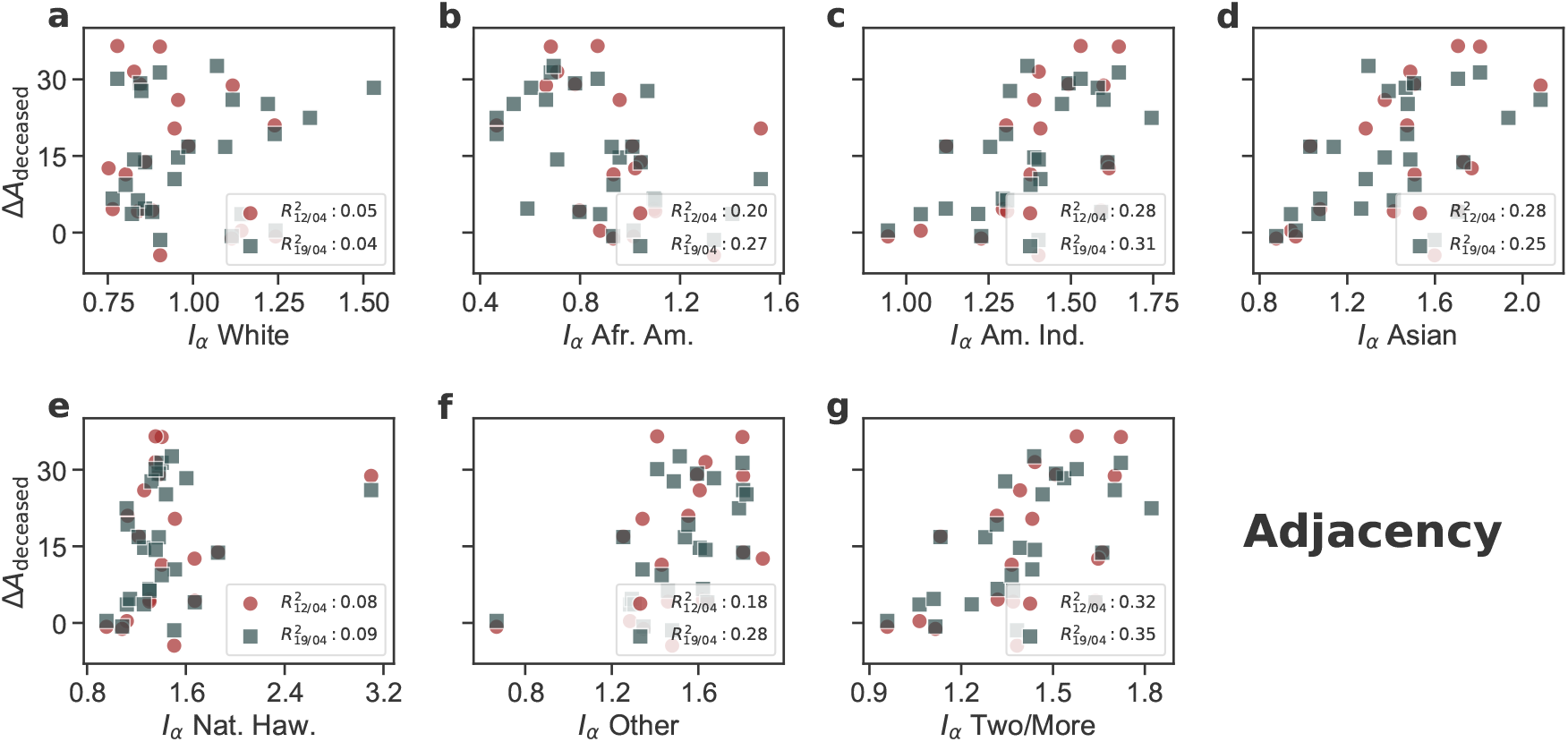
Isolation index of all ethnicities as a function of the deceased rate gap of COVID-19 cases in the African American population considering the adjacency network.

**FIG. S-14.**
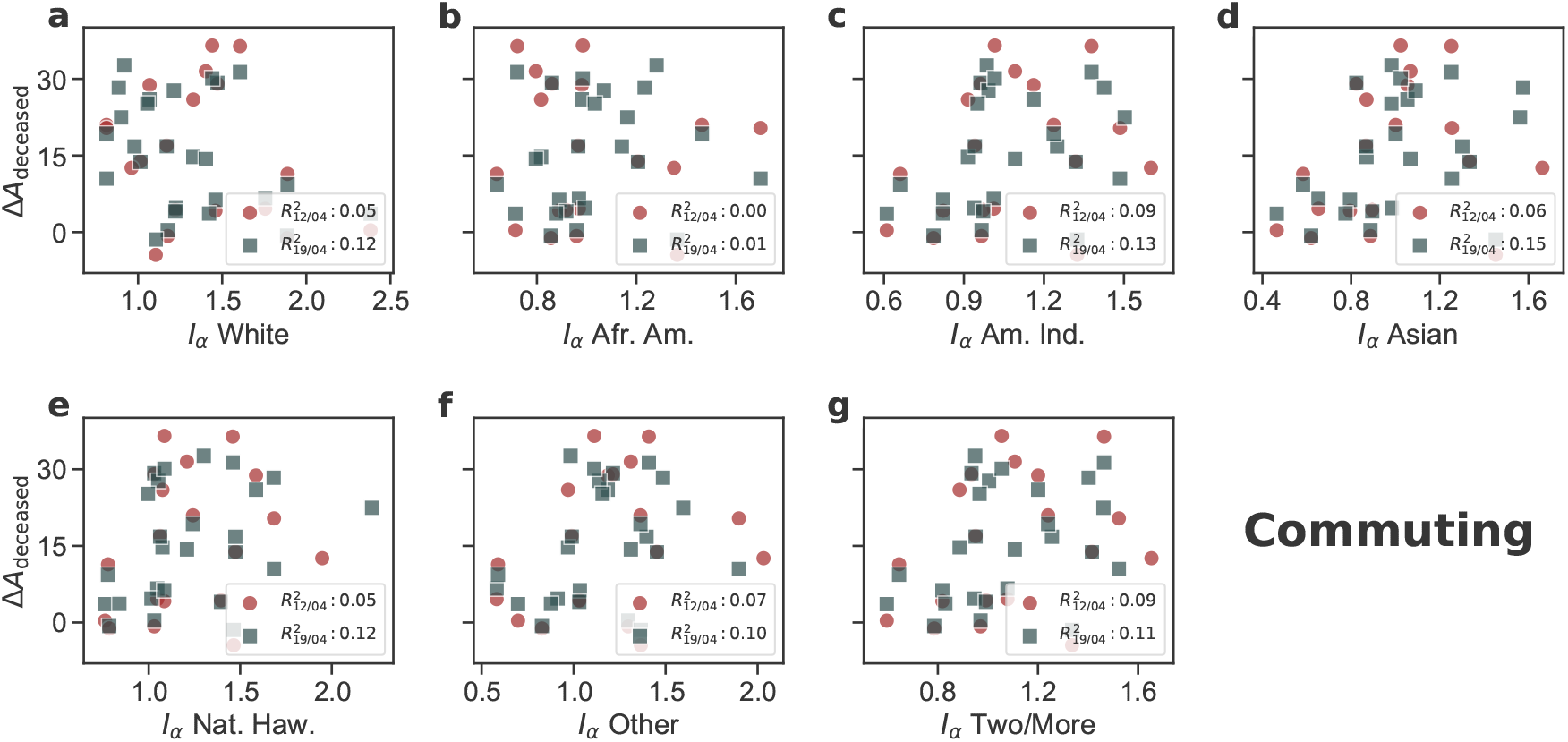
Isolation index of all ethnicities as a function of the deceased rate gap of COVID-19 cases in the African American population considering the commute network. There is no significant correlation for any of the ethnicities.

## Appendix C Appendix C: Local segregation maps through CMFPT and CCT and spatial correlation

In the main manuscript, we show the values for the local segregation indices *ξ* and*Ψ* for Chicago and Los Angeles showing that there were significant differences on their spatial distribution as well as in their maximum values. Here we provide also results for Detroit and Houston to show that again there are significant differences. In this case, Detroit is the most populated city in Michigan, which is one of the states with highest values in most of the indices considered and Houston is the most populated city in Texas, which is a state with consistent low values in most segregation indices. Regarding the impact of COVID-19 among the African Americans of those states, in Michigan the gap is around 34% in early April and 24% in mid-may. In Texas, instead, the gap is around 2% at the beginning of April and 5% in mid-May.

In the main manuscript and Supplementary Figure S-15 we plot the local measures of segregation in each of the census tracts of Chicago, Los Angeles, Detroit and Houston. Those maps display certain common patterns that we quantify in Supplementary Figure S-16. Therein we have calculated the Kendall *τ*_*k*_ correlation coefficient performing pairwise comparisons of the values for each tract unit. Additionally to the segregation indices, we also compared the values for the ratio of African American population. It is relevant to note that while the value of *τ*_*k*_ for the ratio of African American population and 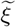 computed in the adjacency graph is around 0.8 for all the four cities studied, there are stronger variations when 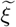 is computed in the commuting graph – i.e., 0.81 in Detroit and 0.55 in Houston – meaning that the effect of commuting in the segregation of African American population can display strong differences across cities and, therefore, mobility offers a different picture of urban segregation.

**FIG. S-15.**
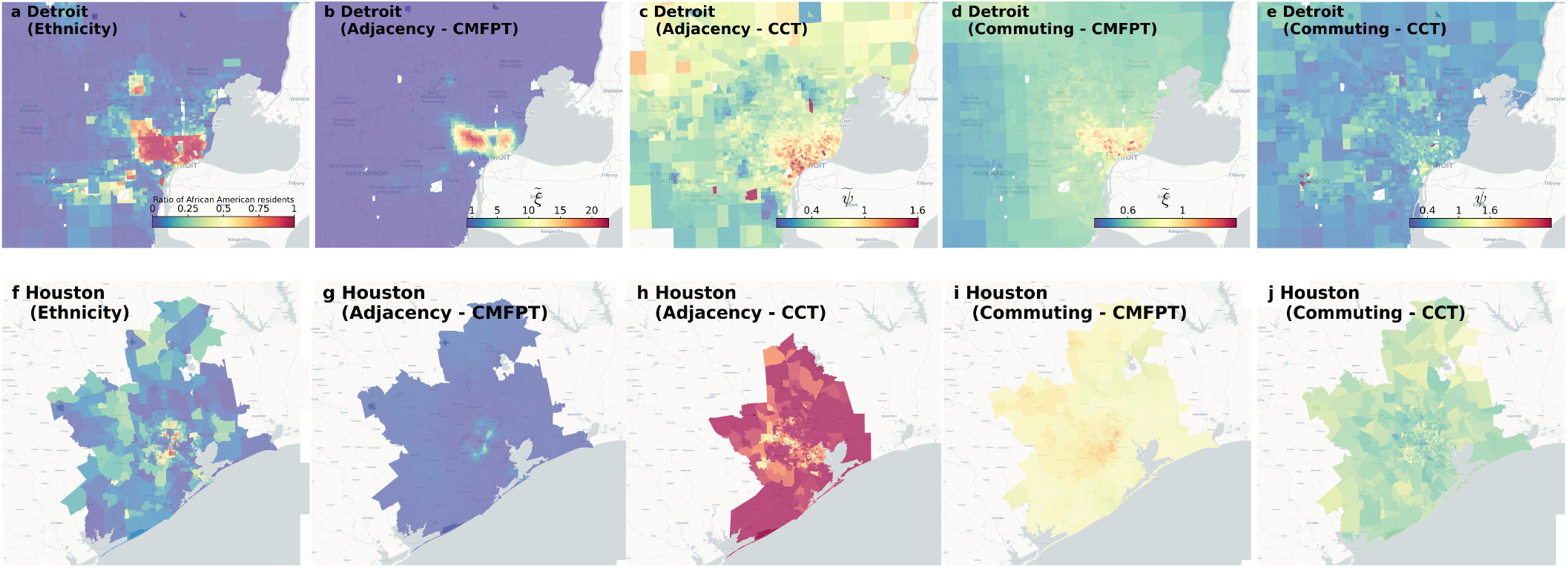
**Maps of local segregation in American cities**. Ratio of African American population and local segregation indices computed with CMFPT and CCT in **a-e** Detroit and **f-j** Houston. For Detroit: **a** Ratio of African American population, **b-c** 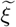 and 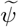 computed over the adjacency graph and **d-e** 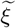 and 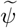 computed over the commuting graph. For Houston: **f** Ratio of African American population, **g-h** 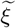 and 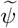 computed over the adjacency graph and **i-j** 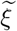 and 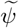 computed over the commuting graph.

## Appendix D Temporal analysis of correlations with segregation indices and other socioeconomic indicators

### Statistical analysis of the COVID-19 incidence data

In this section, we provide the temporal evolution of correlations between the difference in the percentage of COVID-19 incidence among African Americans and other segregation indices. First of all, Supplementary Figure S-17 shows the number of states included in each of the temporal snapshots. As can be seen, it increases with time yet already in the first temporal snapshots there are almost 20 of them. It is important to note that by mid-April, the US reached the first peak of the pandemic.

Additionally to the number of states included in the analysis we also observe significant changes in the values across time for the different states analysed. We provide in Supplementary Figures S-18 and S-19 the evolution of the difference in percentage of infected and deceased African Americans. States are split in quartiles of the distribution of the percentage of African Americans among the overall population. While the average seems almost stable in most of the quartiles this is more a product of compensating changes than of stability in the values for a single state. For instance, in the first quartile there is a sharp increase in Minnesota compensated by a decrease in DC. On the third quartile, the sharp decrease in Illinois is compensated by the increase in Arkansas. It is also important to note that some states display strong discrepancies between the percentage on deceased and infected as, for instance, Minnesota.

#### Correlations with the ratio of infected African Americans and the difference in percentage and the ratio of deceased African Americans

In the main manuscript, the main variable analysed is the difference in the percentage of African Americans since other factors might influence the deceased and the ratio that can lead to several outliers. In Supplementary Figures S-20, S-21 and S-22, we show respectively the correlation with the difference in percentage of deceased African Americans, the ratio of infected and the ratio of deceased. In the case of the difference in percentage we can see that despite correlations are lower they are stable across time. It is important to note that in the case of deceased individuals other factors like the age or the underlying health conditions might play a significant role. In the case of both ratios, correlations are slightly high in the first snapshots suffer a steeper decrease. Again *C* and *E* computed in the commuting graph seem to outperform the rest of metrics.

**FIG. S-16.**
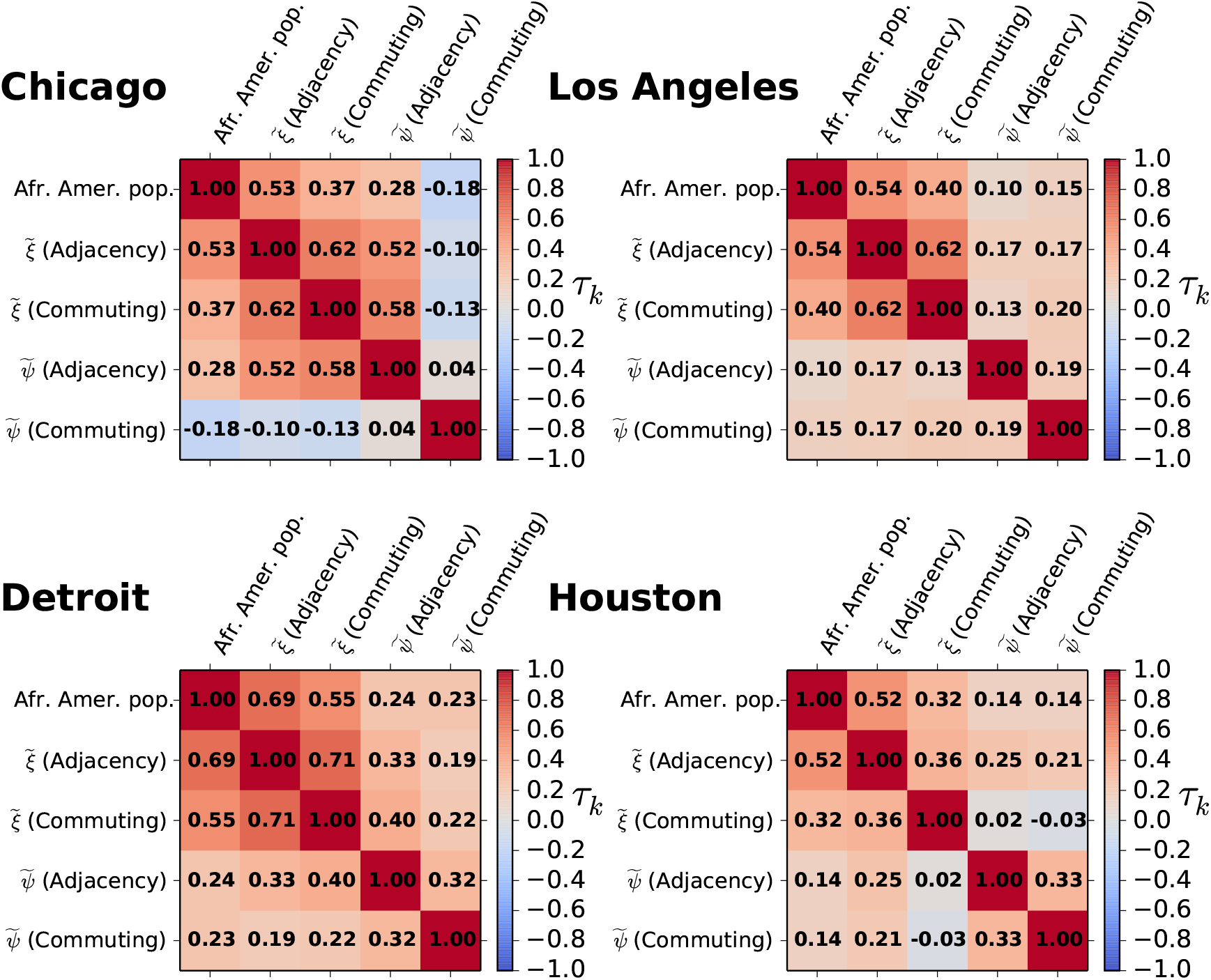
**Correlations between the each of the local metrics of segregation** 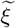 **and** 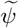 **and the local ratio of African American population**. Correlation between each of the local indices of segregation and the ratio of African American population by census tract as well. On the top row Chicago and Los Angeles and on the bottom row Detroit and Houston.

#### Temporal evolution of correlations with another data set

We also had access to another project that aggregates data on the ethnicity of both infected and deceased African Americans by COVID-19 through three different temporal snapshots 22*/*04*/*2020, 04*/*05*/*2020 and 15*/*05*/*2020 [33]. In Supplementary Figure S-23, we report the correlations in each of the three snapshots for the difference in the percentage of infected African Americans, which are in line with the results obtained for the other data set. Correlations are considerably high and significant for the first stages of the pandemic and decrease with time as the different lock-downs take place. As in the results provided in the main manuscript, those indices computed on the commuting graph seem to provide a better correlation than those computed on the adjacency one. Again those indices connected to the exposure of African Americans only yield significant correlation on the commuting network. The results obtained for the ratio of infected and deceased African Americans as well as the difference on the percentage of deceased are also compatible with those obtained with the previous data set (See Supplementary Figures S-26, S-24 and S-25).

**FIG. S-17.**
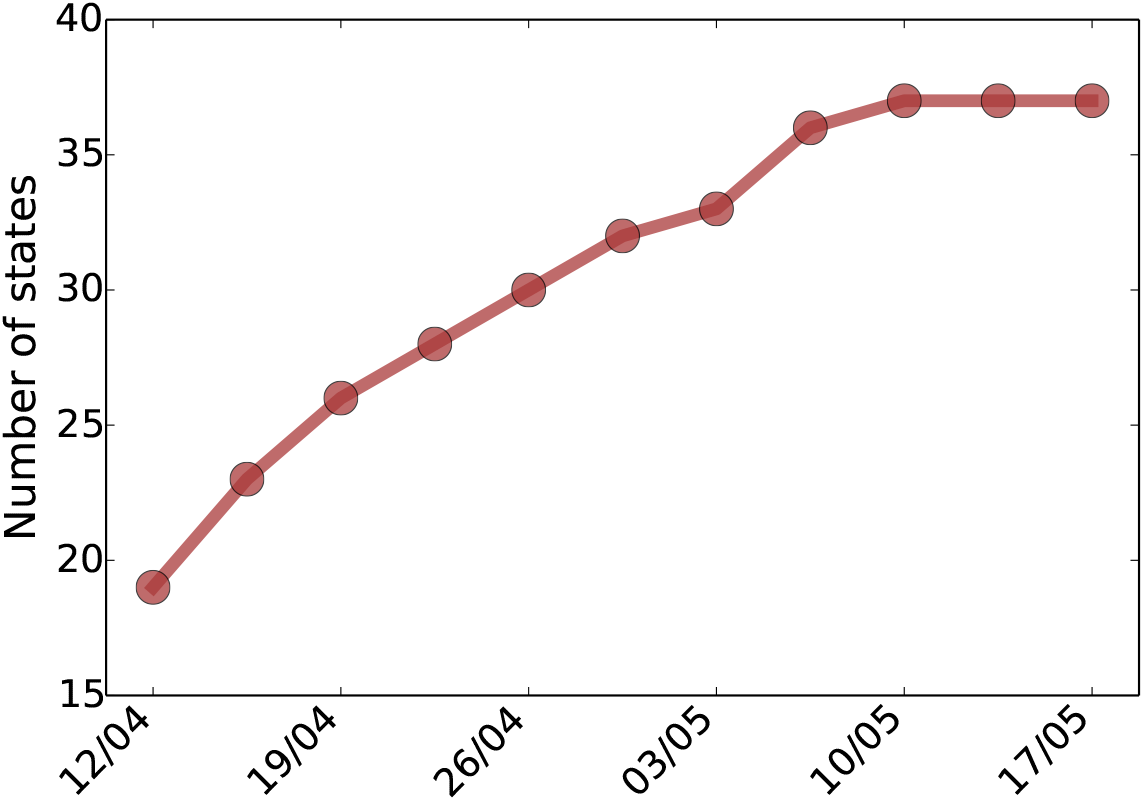
Number of states included in the analysis for each temporal snapshot.

**FIG. S-18.**
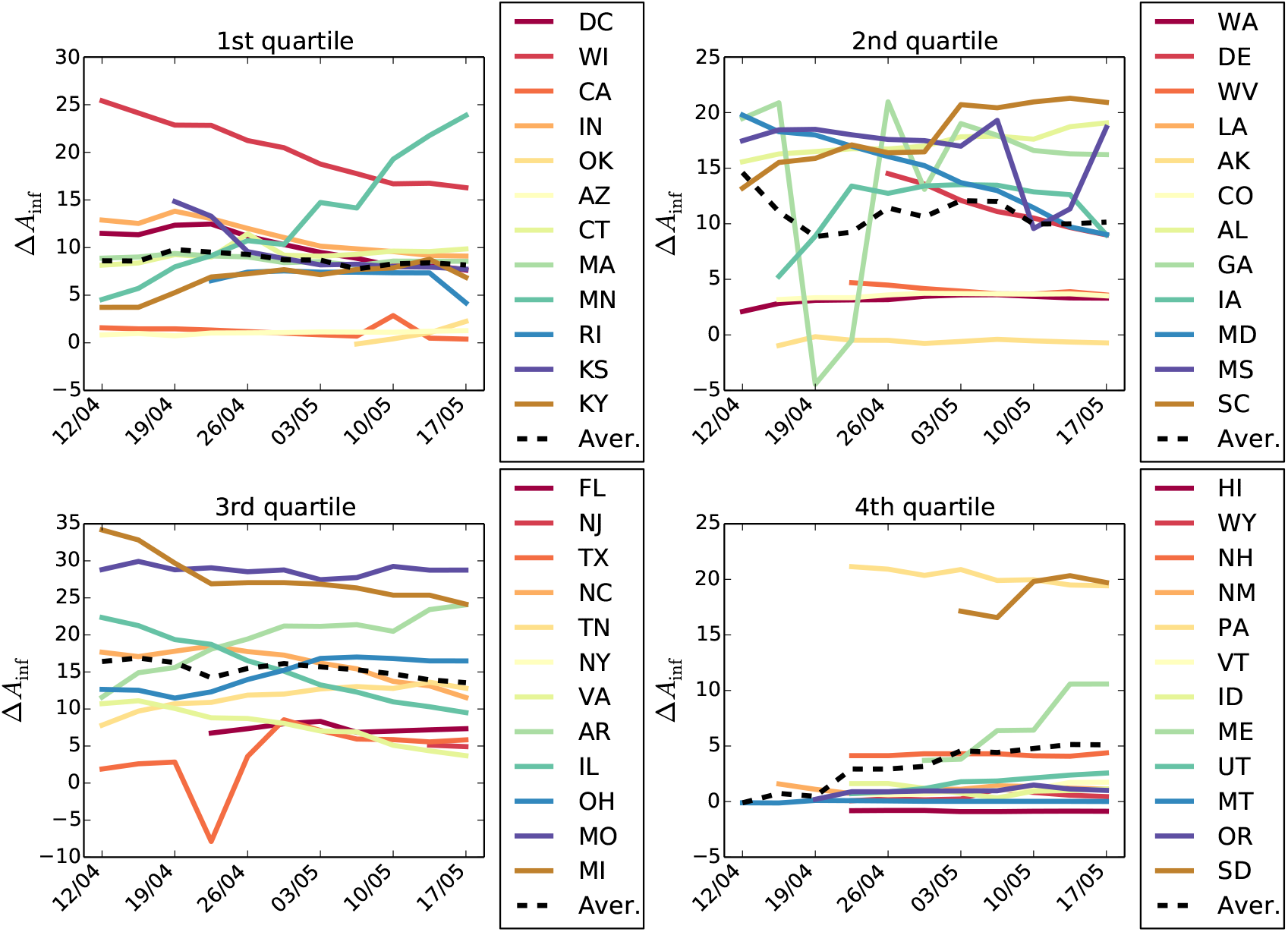
The temporal evolution of the difference in percentage on infected African Americans by state. Each plot represents a quartile of the distribution of percentage of African American population.

**FIG. S-19.**
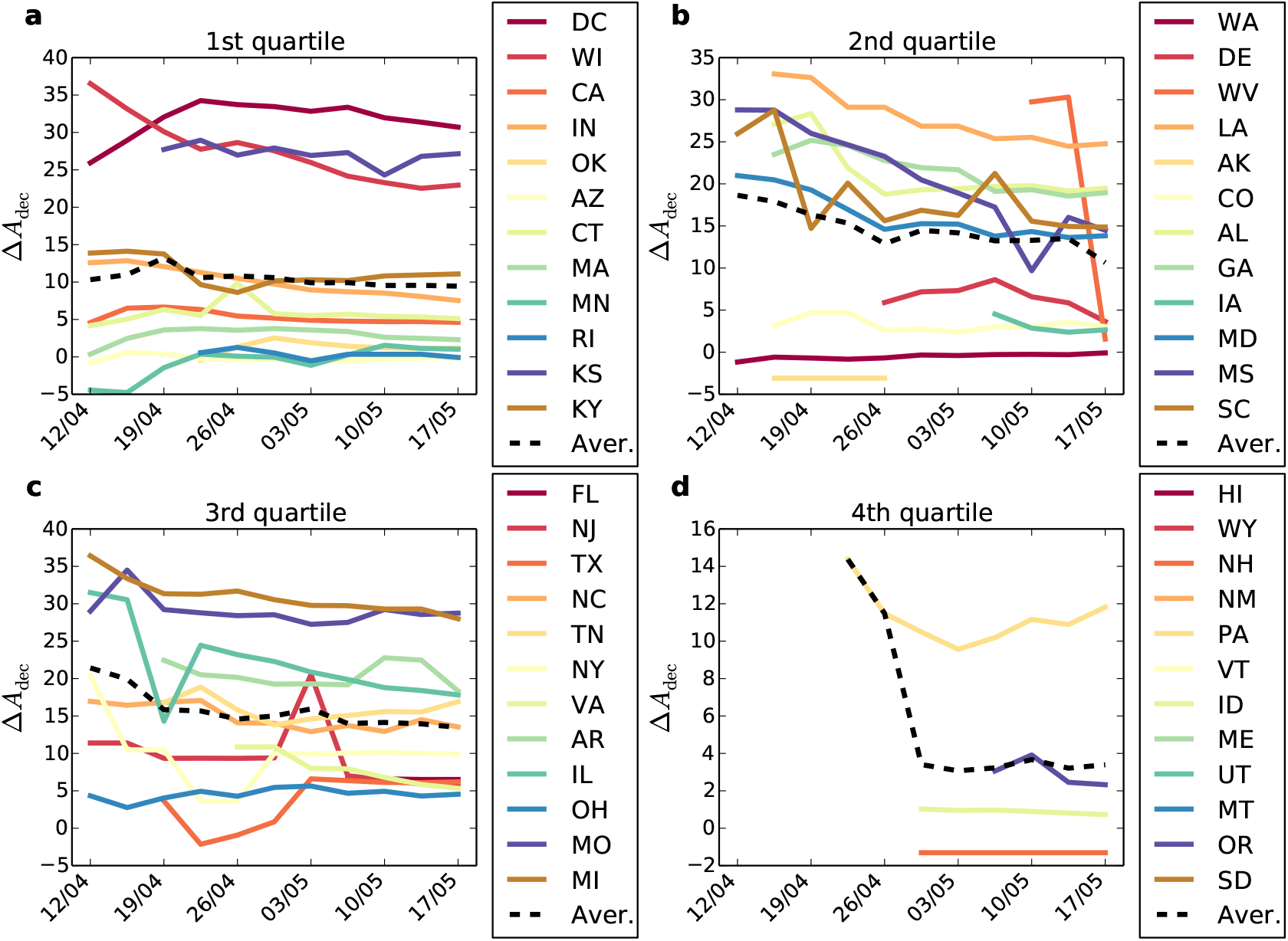
The temporal evolution of the difference in percentage on deceased African Americans by state. Each plot represents a quartile of the distribution of percentage of African American population.

**FIG. S-20.**
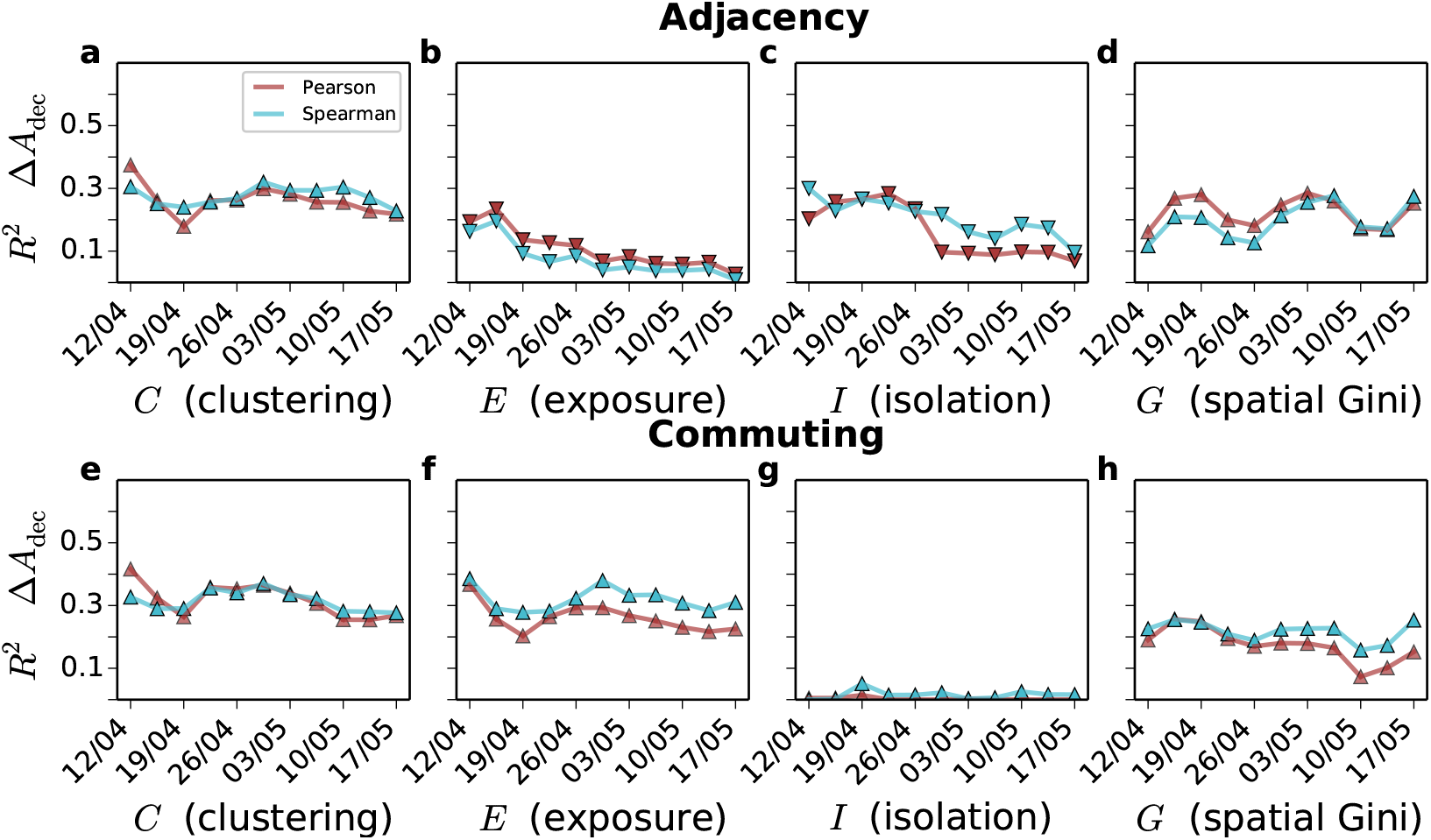
Evolution of the Pearson and Spearman correlation (*R*^2^) found between the difference in percentage of deceased African Americans and the four indices studied in the main manuscript. **a** *C* (clustering), **b** *E* (exposure), **c** *I* (isolation), **d** *G* (spatial Gini). **e-h** Indices computed over the commuting network: **e** *C* (clustering), **f** *E* (exposure), **g** *I* (isolation), **h** *G* (spatial Gini).

**FIG. S-21.**
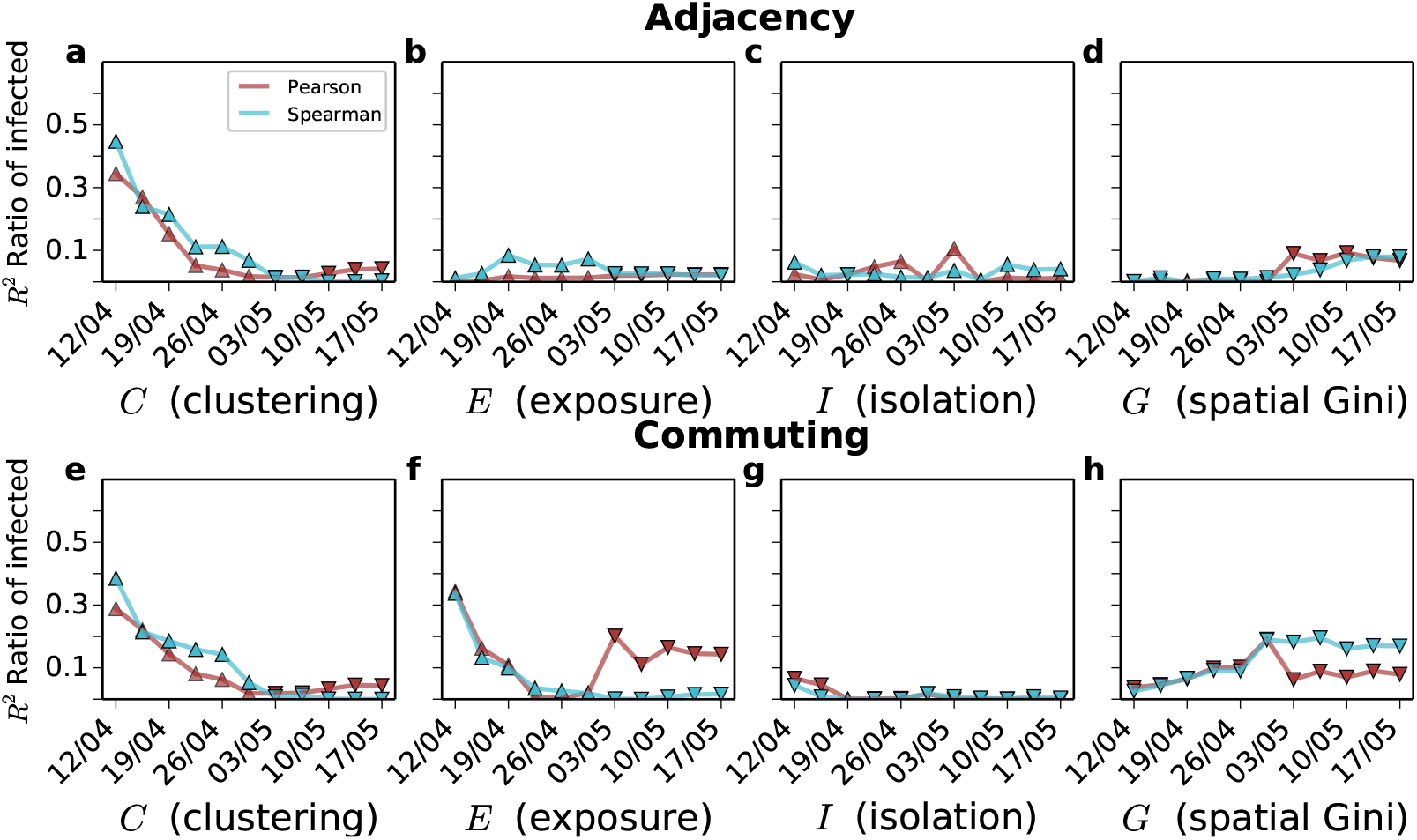
Evolution of the Pearson and Spearman correlation (*R*^2^) found between the ratio of infected African American and each of the indices studied in this work. **a-g** Indices computed over the adjacency network: **a** *C* (clustering), **b** *E* (exposure), **c** *I* (isolation), **d** *G* (spatial Gini). **e-h** Indices computed over the commuting network: **e** *C* (clustering), **f** *E* (exposure), **g** *I* (isolation), **h** *G* (spatial Gini). C (clustering) E (exposure) I (isolation) G (spatial Gini)

**FIG. S-22.**
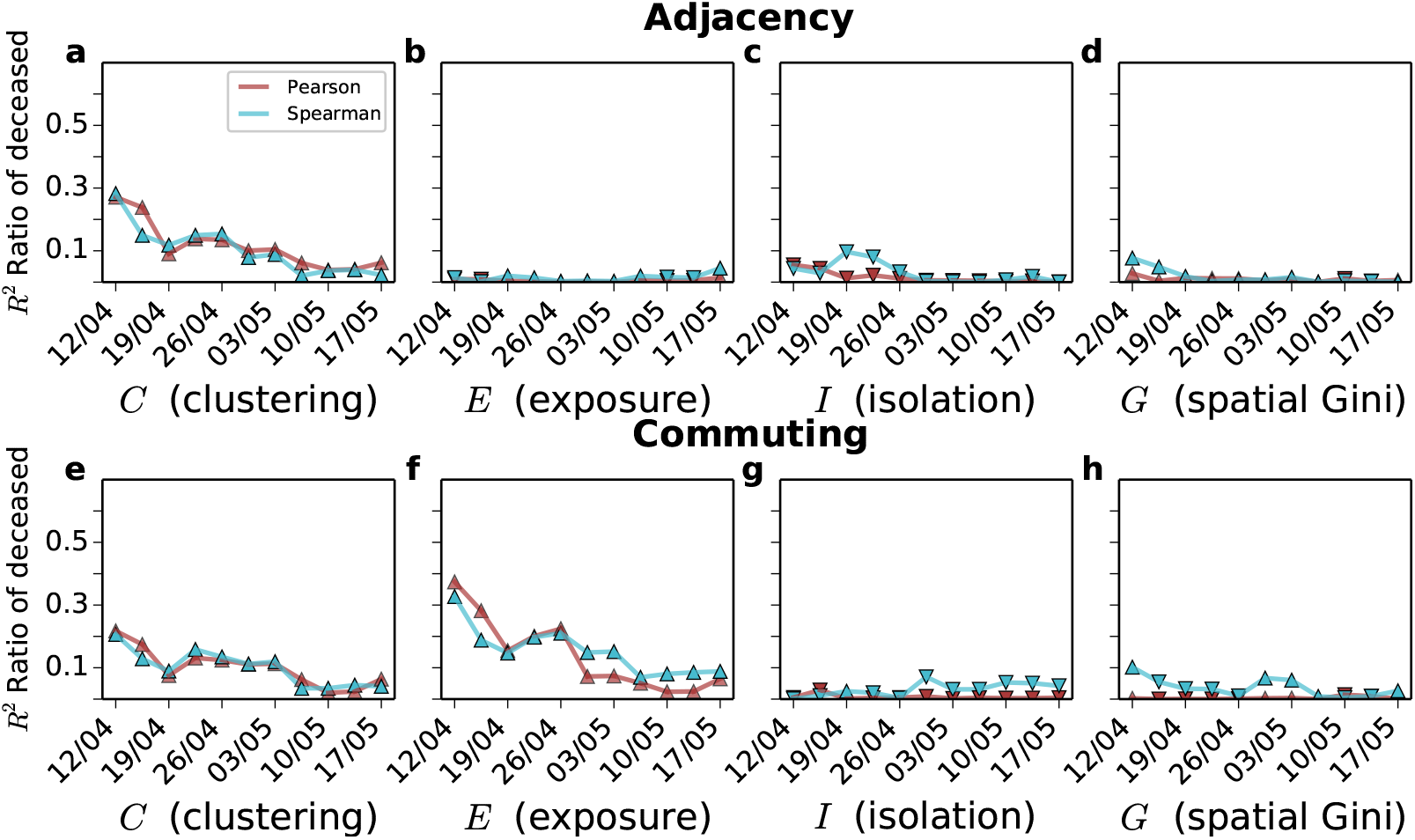
Evolution of the Pearson and Spearman correlation (*R*^2^) found between the ratio of deceased African Americans and the four indices studied in the main manuscript. **a-g** Indices computed over the adjacency network: **a** *C* (clustering), **b** *E* (exposure), **c** *I* (isolation), **d** *G* (spatial Gini). **e-h** Indices computed over the commuting network: **e** *C* (clustering), **f** *E* (exposure), **g** *I* (isolation), **h** *G* (spatial Gini).

**FIG. S-23.**
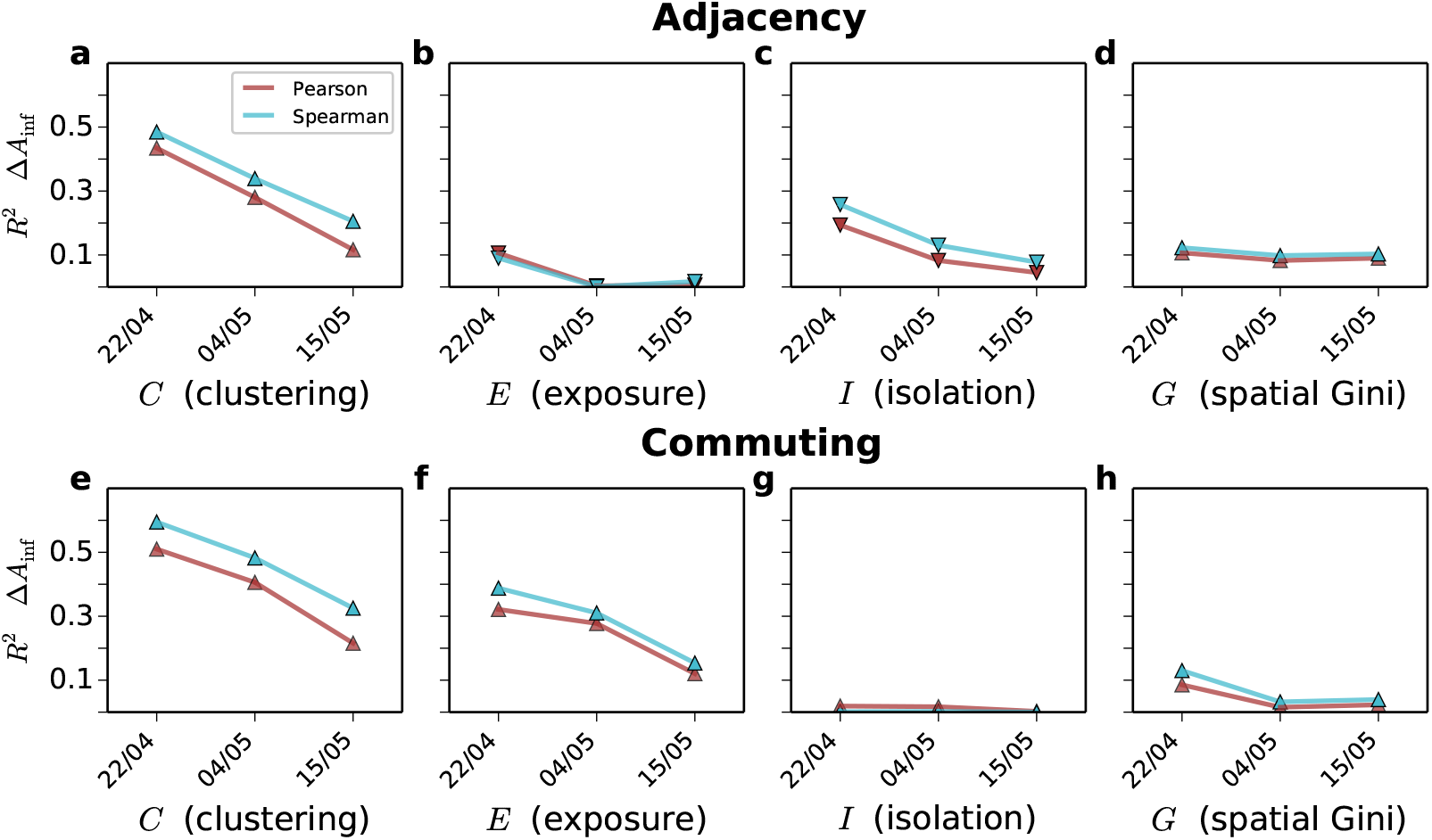
Evolution of the Pearson and Spearman correlation (*R*^2^) found between the difference on the deceased African Americans and the four indices studied in the main manuscript using another data source. **a-g** Indices computed over the adjacency network:**a** *C* (clustering), **b** *E* (exposure), **c** *I* (isolation), **d** *G* (spatial Gini). **e-h** Indices computed over the commuting network: **e** *C* (clustering), **f** *E* (exposure), **g** *I* (isolation), **h** *G* (spatial Gini). The markers indicate the sign of the relation, positive for triangles pointing up and negative for triangles pointing down.

**FIG. S-24.**
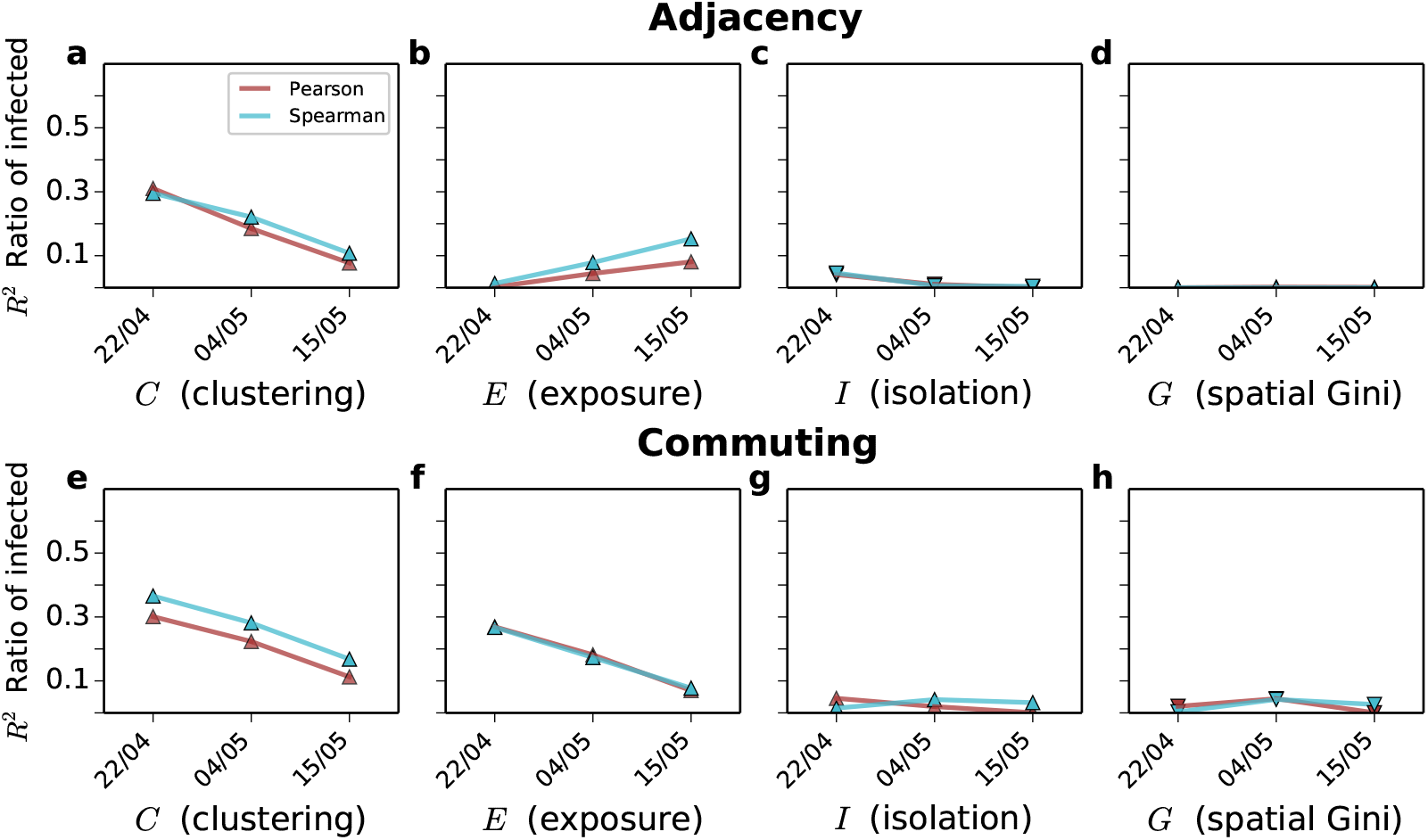
Evolution of the Pearson and Spearman correlation (*R*^2^) found between the difference on the deceased African Americans and the four indices studied in the main manuscript using another data source. **a-g** Indices computed over the adjacency network: **a** *C* (clustering), **b** *E* (exposure), **c** *I* (isolation), **d** *G* (spatial Gini). **e-h** Indices computed over the commuting network: **e** *C* (clustering), **f** *E* (exposure), **g** *I* (isolation), **h** *G* (spatial Gini). The markers indicate the sign of the relation, positive for triangles pointing up and negative for triangles pointing down.

**FIG. S-25.**
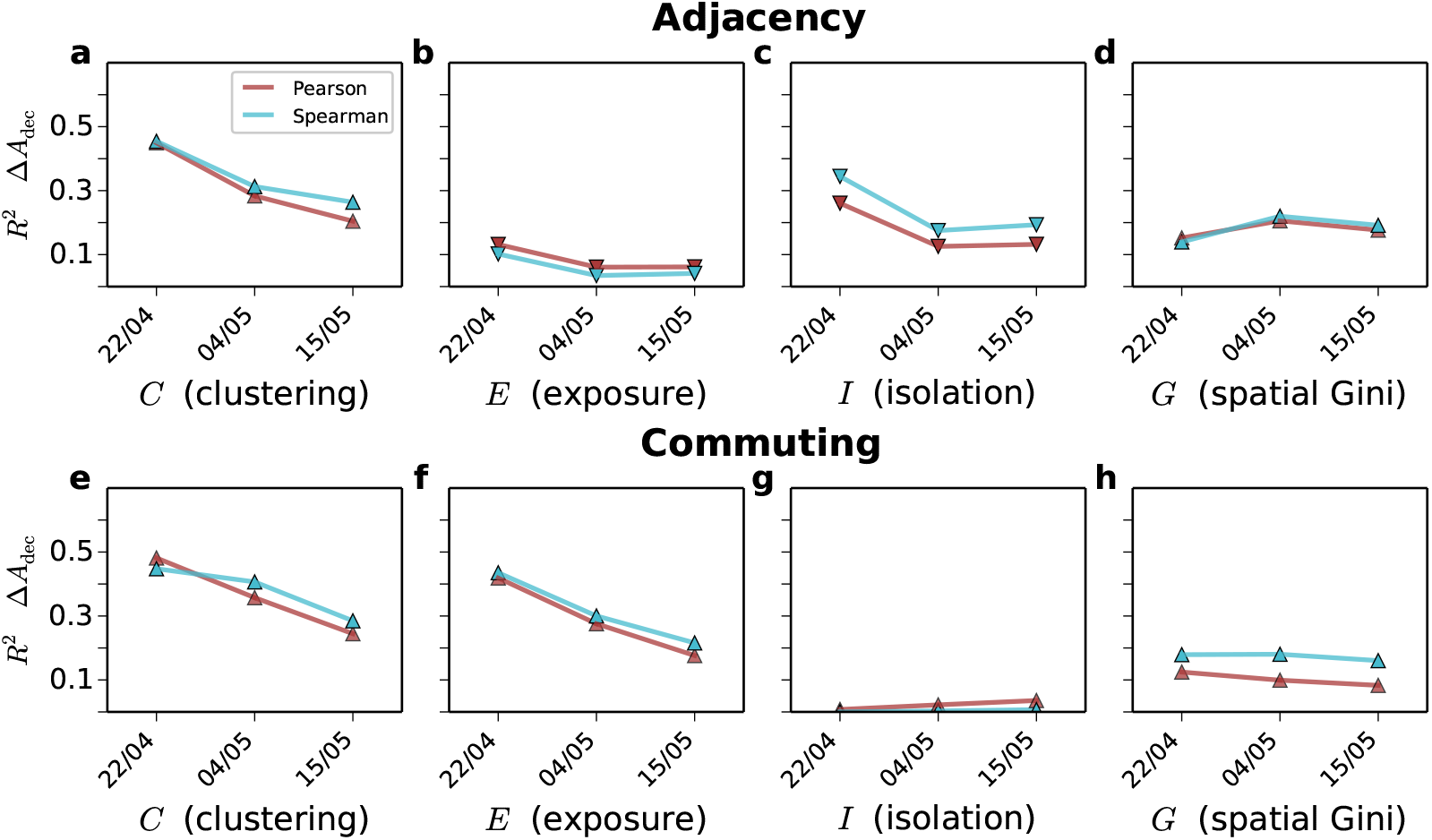
Evolution of the Pearson and Spearman correlation (*R*^2^) found between the ratio of deceased African Americans and the four indices studied in the main manuscript using another data source. **a-g** Indices computed over the adjacency network: **a** *C* (clustering), **b** *E* (exposure), **c** *I* (isolation), **d** *G* (spatial Gini). **e-h** Indices computed over the commuting network: **e** *C* (clustering), **f** *E* (exposure), **g** *I* (isolation), **h** *G* (spatial Gini). The markers indicate the sign of the relation, positive for triangles pointing up and negative for triangles pointing down.

**FIG. S-26.**
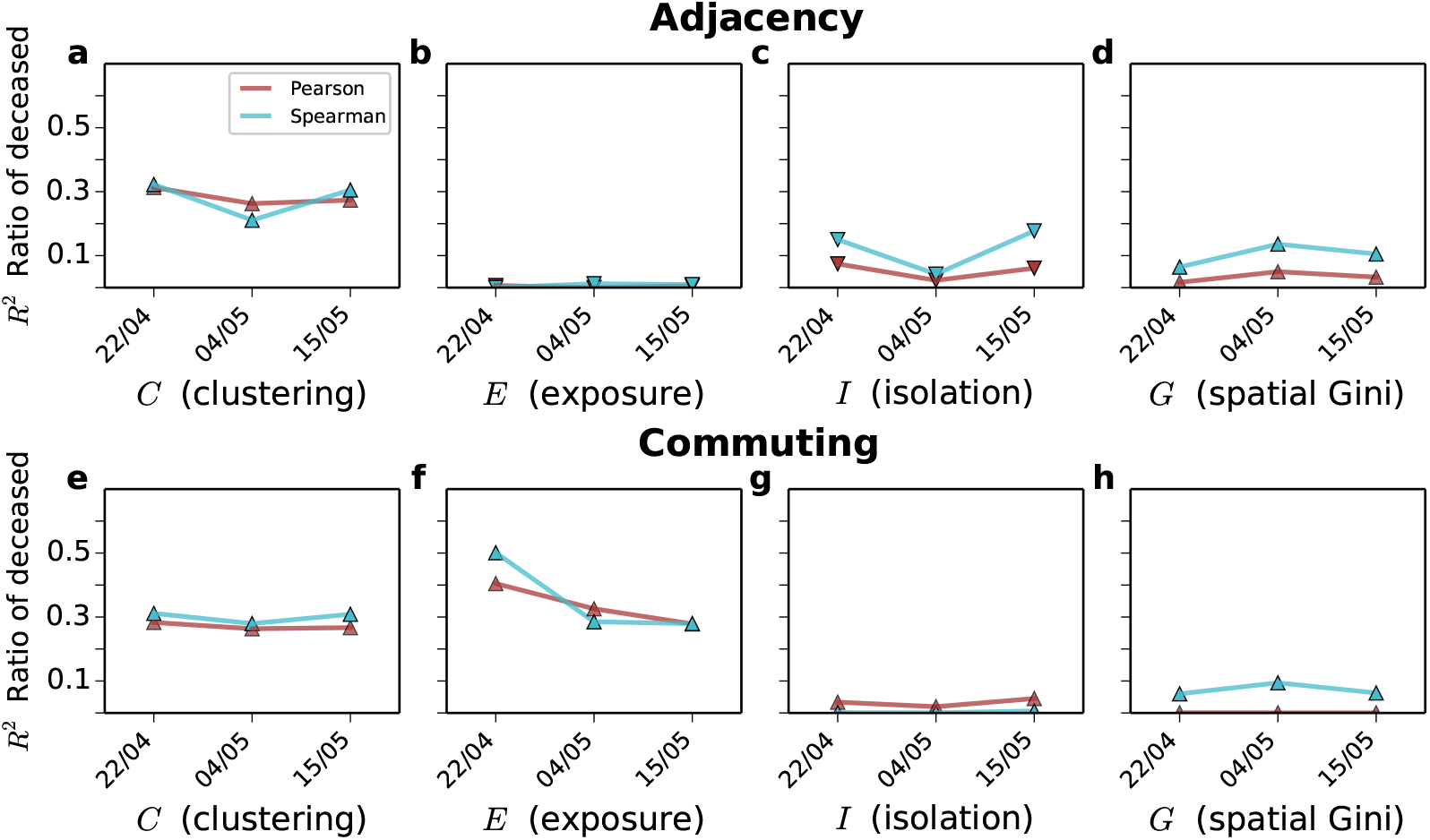
Evolution of the Pearson and Spearman correlation (*R*^2^) found between the ratio of infected African Americans and the four indices studied in the main manuscript using another data source. **a-g** Indices computed over the adjacency network:**a** *C* (clustering), **b** *E* (exposure), **c** *I* (isolation), **d** *G* (spatial Gini). **e-h** Indices computed over the commuting network: **e** *C* (clustering), **f** *E* (exposure), **g** *I* (isolation), **h** *G* (spatial Gini). The markers indicate the sign of the relation, positive for triangles pointing up and negative for triangles pointing down.

#### Formulation of the alternative indices C′ and E′

In the main manuscript we have studied the metrics *C* and *E* which are computed from the elements of the normalised CMFPT 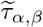. However, there are more potential ways to capture the clustering and exposure of an ethnicity by doing the other calculations from that matrix. Here we propose the two alternative formulations for those two metrics

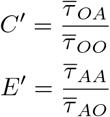

The first quantity comes from the ratio between the time from other ethnicities to African Americans and the time from other ethnicities to others, where higher values correspond more isolated African Americans compared to other ethnicities. The second quantity instead, is the ratio between the time separating African Americans and the time between African Americans and any other ethnicity, where higher values correspond to African Americans more exposed to others than to themselves. The correlation between our alternative proposals and the difference in the percentage of African Americans infected is shown in Supplementary Figure S-27. While correlations are slightly lower, they are still significant. One interesting finding is that *E ′* changes the sign of the correlation when computed over the adjacency graph and the commuting network. Highlighting once again the need of considering mobility to understand the segregation and exposure of ethnicities in urbanscapes.

#### Temporal evolution of correlations with other segregation indices from the literature

We have also studied the correlations between the difference in the percentage of COVID-19 incidence among African Americans and other segregation indices from the literature. First of all, we obtained the segregation index *σ*_*α*_ proposed in [42], which is also based on the movement of random walks is spatial systems and captures the probability that a randomly chosen individual of group *α* meets another individual of the same group, or in this case, ethnicity. Additionally, we also computed Moran’s I, which is a measure of spatial auto-correlation and compares the ethnic composition of neighbourhoods [26]. The correlation of both metrics with the difference in the percentage of infected among African Americans. The evolution of the correlations is shown in Supplementary Figure S-28, where only the Moran index calculated over the adjacency graph seems to display a significant correlation.

For the second metric we have build a matrix of distance between ethnicities similar to the one obtained for 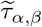 using a measure proposed by [41]. Inspired by the Getis and Ord statistic [49], the metric proposed [41] quantifies for each location *i* the exposure of ethnicity *α* to ethnicity *β* as

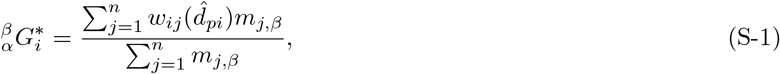

where each *n* is the total number of location in a city, *j* corresponds to each of those locations and *m*_*j,β*_ is the population of ethnicity *β* in location *j*, 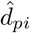 is an estimate of trip length and 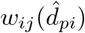 is a function of the distance that is equal to 1 when *d*_*ij*_ *< d*_*pj*_ and 0 otherwise. In the case of the adjacency graph only adjacent pair of tracts were considered whereas in the case of the commuting network only pairs connected by commuting trips were considered. Overall 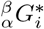 quantify the ratio of population of ethnicity *β* to which the individuals residing in *i* are exposed. In our case we set the threshold *d*_*pj*_ equal to the average commuting distance in each of the cities. Succinctly, 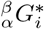 is a value between 0 and 1 that encapsulates the fraction of the population of ethnicity. We average the value of 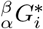 to obtain a distance matrix between ethnicities in each of the cities as

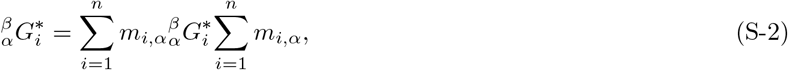

so that we take into account the fraction of population of ethnicity *α* in location *i*. Finally from the matrix 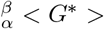 we compute the same exposure and clustering indices computed from 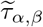 in the main text. Calculating first

**FIG. S-27.**
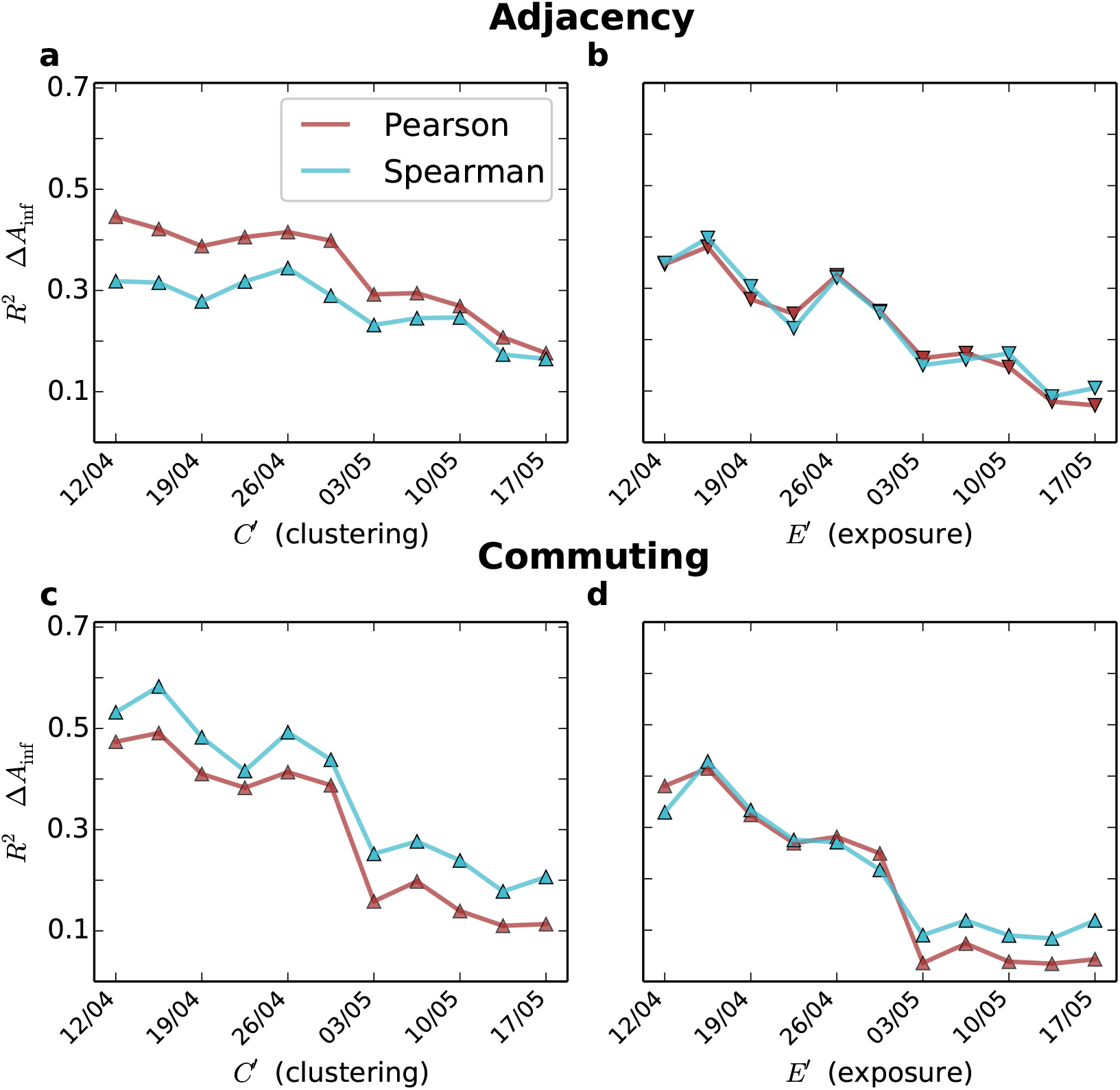
Evolution of the Pearson and Spearman correlation (*R*^2^) found between the difference of infected African Americans and the alternative indices proposed *C′* and *E′*. **a** *C′* (clustering) and **b** *E*^*t*^ (exposure) calculated upon the adjacency network. **c** *C′* (clustering) and **d** *E′* (exposure) calculated upon the commuting network. The markers indicate the sign of the relation, positive for triangles pointing up and negative for triangles pointing down.

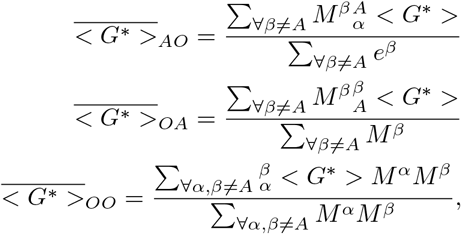

to finally obtain

**FIG. S-28.**
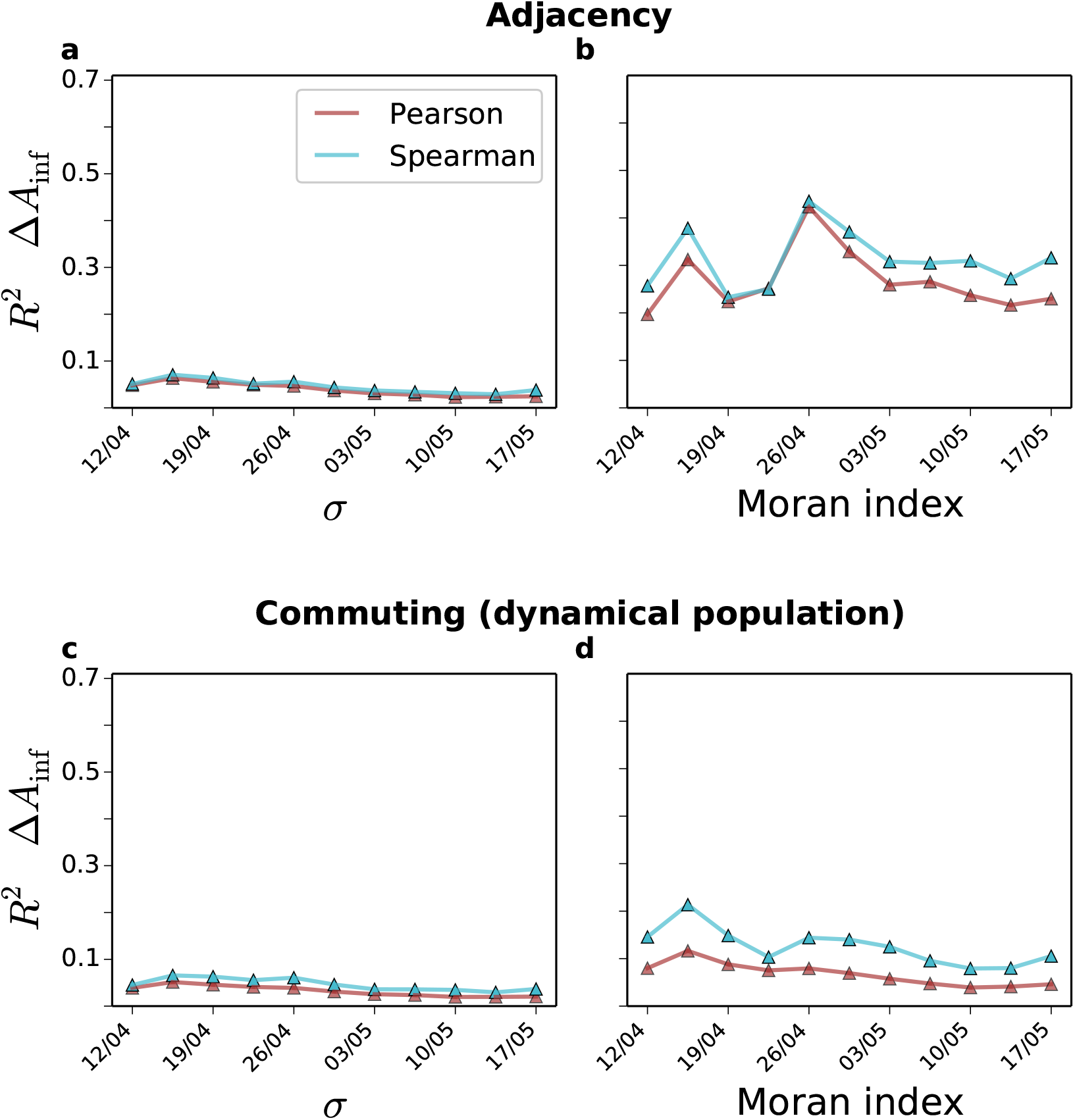
**The temporal evolution of the correlation (***R*^2^**) between the incidence of COVID-19 in African American population and the segregation indices** *σ* **and Moran’s I. a** *σ* and **b** Moran’s I computed over the adjacency graph. **c** *σ* and **d** Moran’s I computed over the commuting graph. The markers indicate the sign of the relation, positive for triangles pointing up and negative for triangles pointing down.

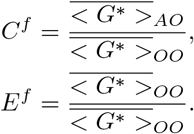

Additionally to the calculation of the indices in the adjacency and the commuting network with dynamical population we also computed it with the residential population and the commuting network to investigate the role played by the dynamical population. As can be seen in Supplementary Figure S-29, significant correlations appear with all indices yet the higher ones are with the exposure index especially when computed on the commuting network with dynamical population. Correlations are, however, lower and less stable than those obtained in the main manuscript. Overall, it is important to note that none of the additional segregation metrics we have studied in this section is more informative than the ones we proposed on the main manuscript based on CMFPT and CCT. Moreover, the use of the dynamical population together with the commuting network seems to improve some of the indices.

**FIG. S-29.**
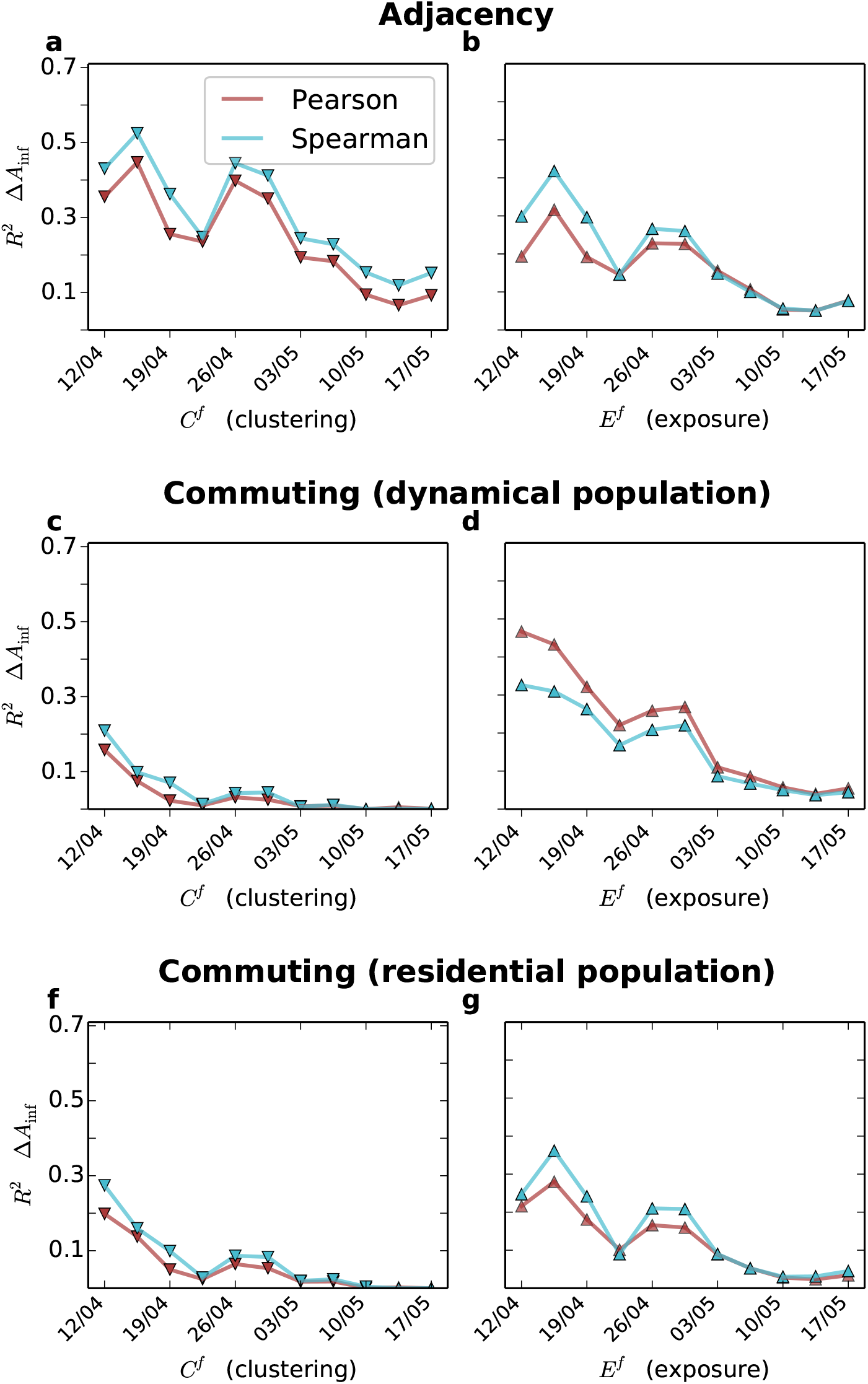
**Correlations between the incidence of COVID-19 in African American population and the clustering and exposure indices computed from the** *G*^*∗*^ **statistic proposed in [41]. a**,**b** Correlation with the clustering *C*^*f*^ and exposure *E*^*f*^ indices computed over the adjacency graph. **c**,**d** Correlation with the clustering *C*^*f*^ and exposure *E*^*f*^ indices computed over the commuting graph when the dynamical population is incorporated. **e**,**f** Correlation with the clustering *C*^*f*^ and exposure *E*^*f*^ indices computed over the commuting graph when only the residential population is incorporated. The markers indicate the sign of the relation, positive for triangles pointing up and negative for triangles pointing down.

#### The relation between the incidence of COVID-19 in African Americans and socio-economic indicators

We present in this section the correlations between a set of socio-economic indicators and the incidence of COVID-19 in African Americans. For the sake of brevity, we focus here only on the data set used in the main manuscript as well as in the difference in the percentage of infections which is the case where correlations are higher. The set of indicators we have studied are the median household income, the percentage of the population below the poverty level, the percentage of insured and uninsured African Americans, the usage of public transportation by both African Americans and the overall population, the percentage of African American population in a state, the average commuting distance and the ratio between the average commuting distance of African Americans and the overall population. All of the metrics are provided at the level of the African American population and the results are shown in Supplementary Figure S-30. The median household income, the percentage of the population below the poverty level, the percentage of insured and uninsured African Americans were obtained from the 2018 American Community Survey elaborated by the U.S. Census Bureau [34]. Most of the variables yield low or very low correlations except for the usage of public transportation by African Americans. Economic indicators such as median income or percentage of poverty seem to slightly correlate with the incidence of COVID-19, which could because because a more deprived African American community puts them in a more risky situation. Regarding the health indicators related to the degree of insurance of African Americans, it seems there is no direct relation with the number of infected. Not so surprising results since we are analysing the percentage of infected and, therefore, the fact of having insurance might not change significantly the risk of getting the illness. Finally, the usage of public transportation seems to play a crucial role in the spread of the disease, especially if we compare the use done by the African American population and the overall population where no correlation appears. The fact that African Americans use more public transportation might put them on a more dangerous position as well as might a reflection of their economic status. Moreover, it could happen that in those cities in which African Americans are more segregated they also have to use more the public transportation.

Additionally to those socio-economic variables we also tested if the overall African American population can also be used as a proxy for the difference in percentage. We also computed on our commuting networks the average commuting distance of the African American population as well as the ratio with the commuting distance of the overall population. As displayed in Supplementary Figure S-31, the overall percentage of African American population seems to be related to the difference in the percentage of infected. However, there is a striking difference between the Pearson and the Spearman correlation coefficients, which means that the rank is more or less conserved yet there are strong outliers. In other words, a state with more percentage of African American population will more easily have a higher difference on the infected yet the population does not align the points in a straight trend. Regarding the mobility indicators, none of them yields a significant correlation, meaning that it is not so relevant how far African Americans travel and where they travel and whom they meet.

**FIG. S-30.**
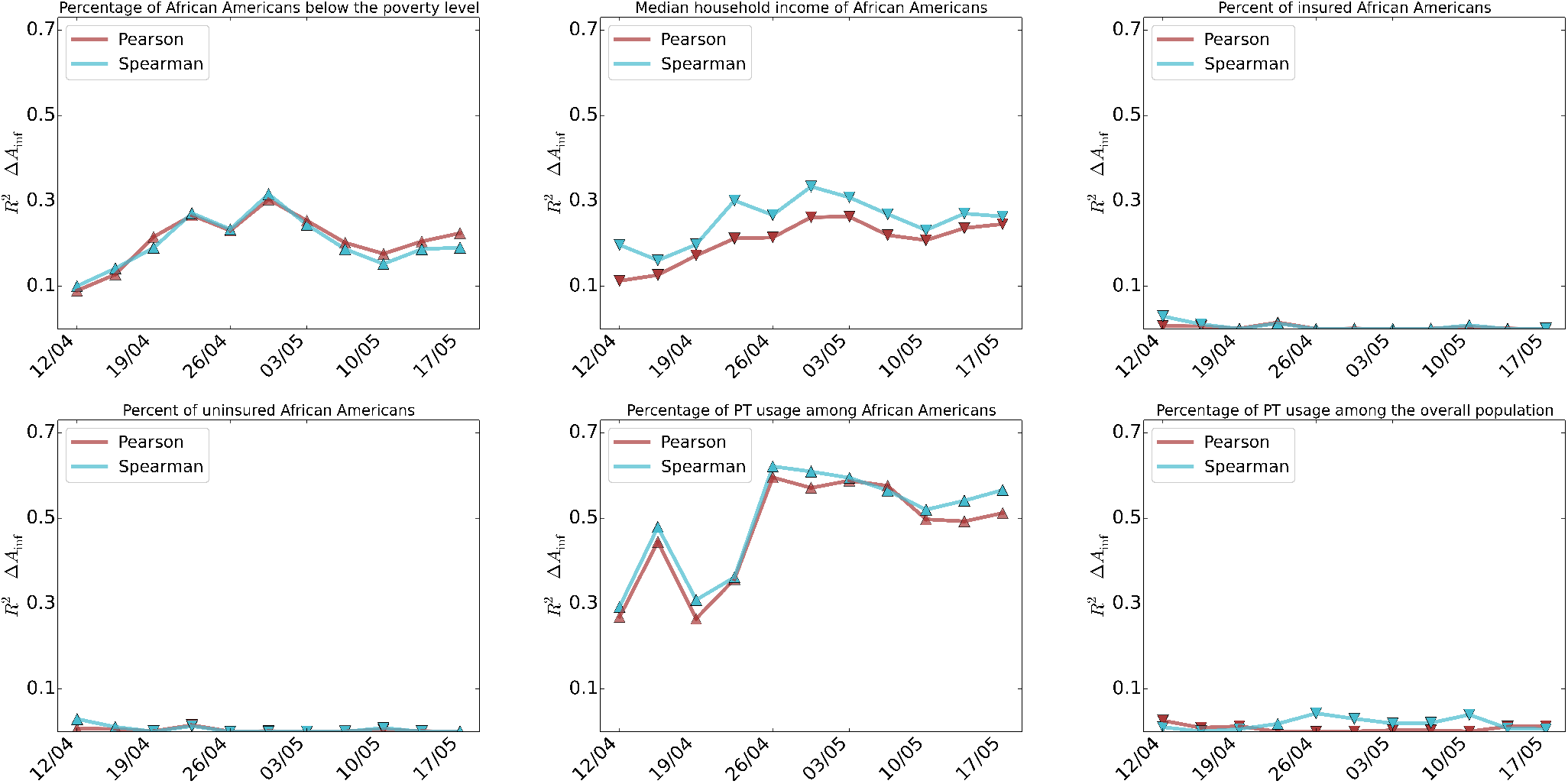
**The temporal evolution of Pearson and Spearman correlations (***R*^2^**) between the incidence of COVID-19 in African American population and a set of socio-economic indicators**. On the top row and from left to right we have the median household income of African Americans, the percentage of African Americans below the poverty level and the percent of insured African Americans. On the bottom row and from left to right there is the percent of uninsured African Americans, the percentage of use of public transportation among African Americans and the percentage of use among the overall population. The markers indicate the sign of the relation, positive for triangles pointing up and negative for triangles pointing down.

**FIG. S-31.**
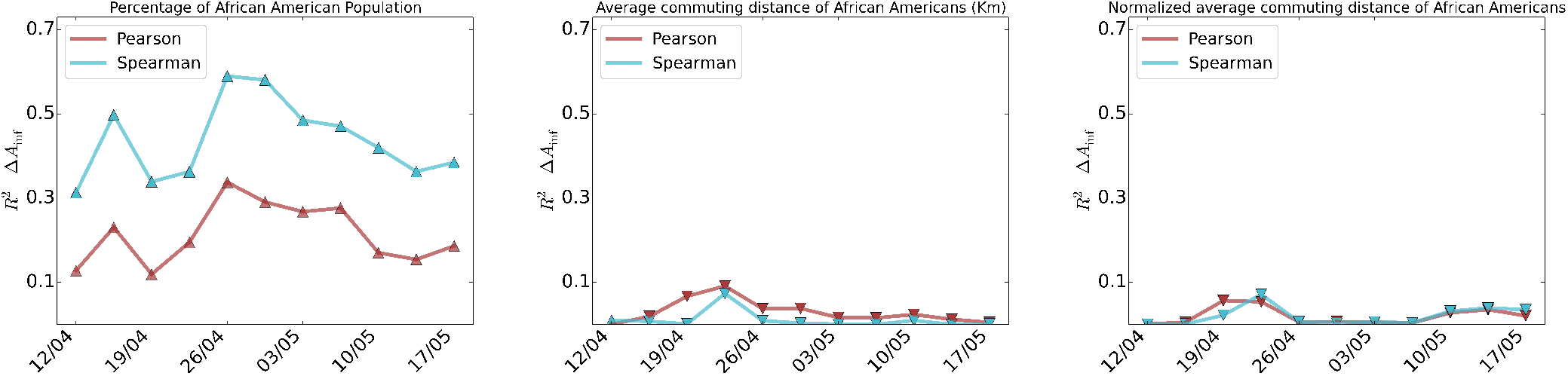
**Correlations between the incidence of COVID-19 in African American population and a set of population and mobility indicators**. From left to right the temporal evolution of the Pearson and Spearman *R*^2^ for the percentage of African Americans among the population, the average commuting distance of African Americans and its ratio with the commuting distance of the overall population. The markers indicate the sign of the relation, positive for triangles pointing up and negative for triangles pointing down.

## Notes

### Competing Interest Statement

The authors have declared no competing interest.

